# A novel automated participant-recorded dietary data collection method using low-cost mobile phones and Interactive Voice Response (IVR) with low-literacy women: a validation study in rural Uganda

**DOI:** 10.1101/2025.08.19.25333742

**Authors:** Lydia O’Meara, Kate Wellard, Joweria Nambooze, Patrick Ongora, Paula Dominguez-Salas, Elaine Ferguson

## Abstract

**Background:** Dietary data gaps limit the effectiveness of food policies and programmes in low-and middle-income countries (LMICs). Automated mobile phone-based tools could fill data gaps at reduced time and cost compared with face-to-face methods, especially for high-frequency dietary quality monitoring in resource-constrained environments.

**Objective:** This study assessed the validity of automated participant-recorded Interactive Voice Response (IVR) 24-hour dietary recalls to assess dietary quality amongst marginalised rural women in a sub-Sahara African context, against same-day gold standard observed weighed food records (WFR).

**Methods:** Automated IVR with push-button response on basic mobile phones collected semi-quantitative list-based 24-hour dietary recalls from 156 randomly selected women in rural Northern Uganda during the wet season. Inter-method agreement was assessed by comparing mean women’s dietary diversity scores (WDDS), the percentage achieving minimum DDS for women (MDD-W), and consumption of unhealthy foods and beverages.

**Results:** Most women (74.4%) completed the IVR. Compared with the WFR, agreement for the IVR was moderate for MDD-W (21.6% vs. 15.5%; *kappa*=0.52; area under the curve=0.80), mean WDDS (3.3 vs. 3.5; weighted *kappa*=0.41), and unhealthy food (34.5% vs 23.3%; *kappa*=0.44) and beverage consumption (32.8% vs 31.9%; *kappa*=0.43).

**Conclusion:** This is the first study to validate the use of IVR via basic mobile phones to collect dietary data to estimate population-level MDD-W, WDDS and percentage consuming unhealthy foods and beverages amongst marginalised rural women in sub-Saharan Africa. With provision of short participant training, results indicate this innovative automated method can be used in place of enumerator-administered methods for monitoring key dietary quality indicators widely used in LMICs with low-literate, rural women in Uganda.

**Highlights:**

- This is the first study to validate the use of Interactive Voice Response (IVR) to collect dietary data from digitally marginalised rural women in a sub-Saharan African context, for estimating key international dietary quality indicators widely used in LMICs - MDD-W, WDDS, and the percentage consuming unhealthy foods and beverages
- This innovative method can be used in place of conventional enumerator-administered methods, after contextualisation and training participants on its use
- Such methods can help fill critical data gaps on dietary quality with low-literate women in resource-scarce settings

Figure 0:
Graphical abstract

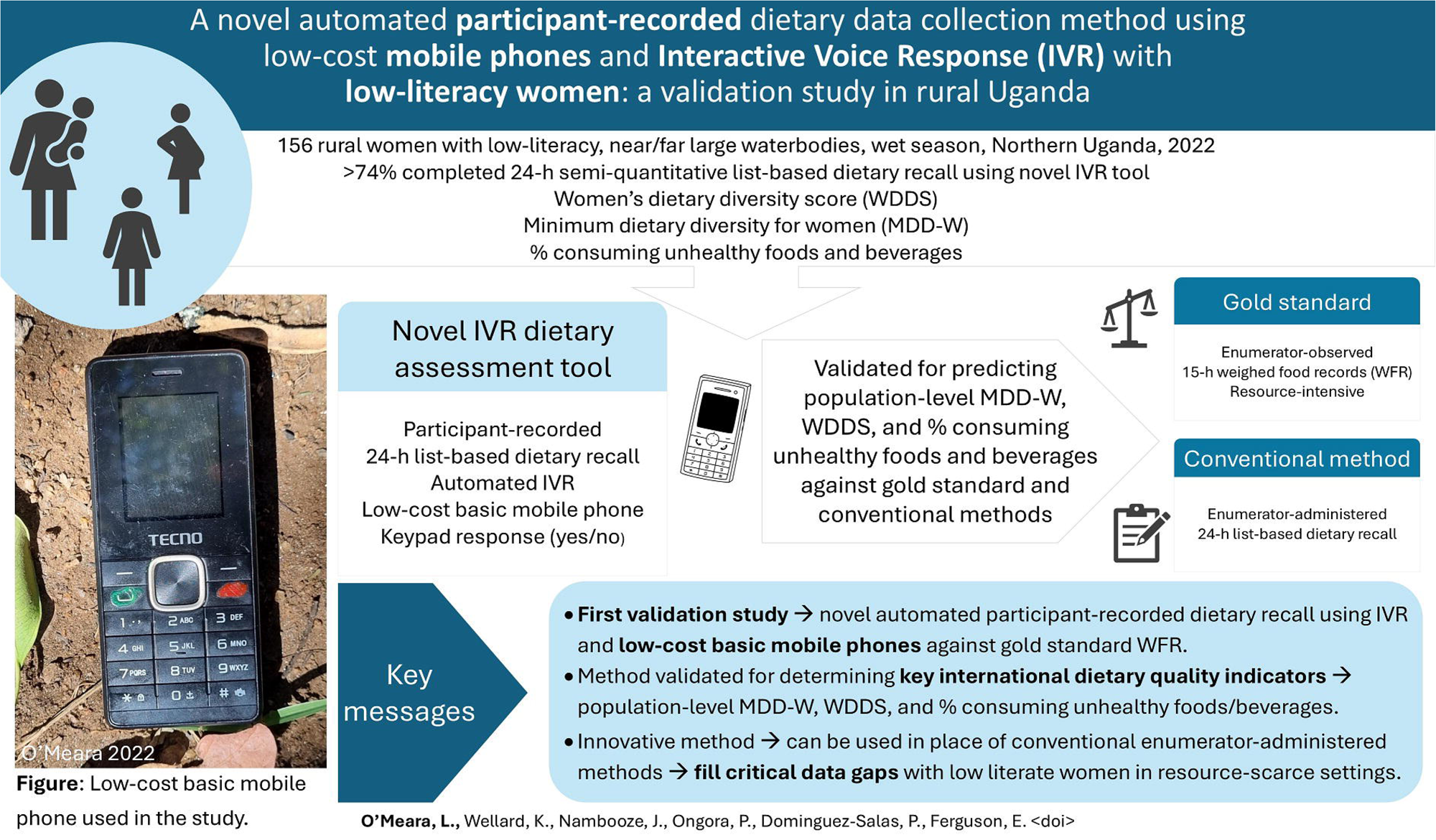

## Introduction

Low dietary quality and malnutrition are the single largest contributors to morbidity and mortality worldwide (IHME, 2025), particularly among women and children in low- and middle-income countries (LMICs) (FAO/IFAD/UNICEF/WFP/WHO, 2024). Dietary quality is a critical indicator for tracking progress toward global development goals, including improving nutrition and promoting sustainable food systems (Fanzo et al., 2021; FAO, 2025). Effective policies and interventions are hindered by a paucity of dietary data (Coates et al., 2017; Schneider et al., 2023), exacerbated by costs and challenges in monitoring marginalised and vulnerable populations. Conventional enumerator-administered methods face barriers such as geographical remoteness due to distance or poor infrastructure (FAO, 2018), population movement (Jensen et al., 2023), and high resource requirements (Vecino-Ortiz et al., 2021). Cyclic wet seasons, often known as ‘hungry’ periods, necessitate close monitoring of food security and dietary quality; however, face-to-face data collection during heavy rains can be resource intensive or infeasible (FAO, 2018). Such challenges are most acute during crises—such as epidemics and pandemics, severe weather events, and conflicts—when dietary risks peak (O’Meara et al., 2024, 2025), but data collection becomes logistically constrained (Picchioni et al., 2022). With the projected intensification of climate change, conflict, and infectious disease outbreaks threatening the food security of marginalised communities (FAO/IFAD/UNICEF/WFP/WHO, 2024), innovative participant-recorded dietary assessment tools will be valuable for filling dietary data gaps (De Bruyn et al., 2019; Jensen et al., 2023) and enhancing real-time monitoring with limited resources (Al Kibria et al., 2023; Manners et al., 2022; Vecino-Ortiz et al., 2021). Such cost-effective and rapidly collected dietary data, if accurate, would enable timely and targeted food security and nutrition responses in LMICs, especially among the most marginalised where the burden of malnutrition remains disproportionately high (FAO/IFAD/UNICEF/WFP/WHO, 2024).

Worldwide, telephones are the most widely used and fastest growing, two-way communication channel in LMICs, with approximately 80% of women in LMICs owning a mobile phone (GSMA, 2023). Whilst female mobile phone ownership can be much lower in rural low-resource settings such as remote sub-Saharan African contexts (Wesolowski et al., 2012), mobile phone network coverage and availability of low-cost basic mobile phones is increasing rapidly in LMICs (GSMA, 2023). During the worldwide movement restrictions of the Covid-19 pandemic, researchers relied on telephone-(Picchioni et al., 2022) and web-based surveys (O’Meara et al., 2022) to assess dietary changes in LMICs. However, in LMICs, existing mobile phone approaches for collecting semi-quantitative dietary diversity scores are enumerator-reliant (Assefa et al., 2022; Lamanna et al., 2019), whilst text-message (Manners et al., 2025) and web-based surveys (O’Meara et al., 2022) privilege high literate groups.

Two reviews highlight a lack of validated digital tools for dietary monitoring in LMICs, with a paucity of studies validating dietary diversity data collection using automated participant-recorded mobile phone methods against gold standards (Greenleaf et al., 2017; Sparling et al., 2019). Three recent studies assessed the potential for collecting participant-recorded dietary data from women in LMICs using mobile phones (Manners et al., 2025; Jensen et al., 2023; Folson et al., 2023). One study explored the validity of using text-message based surveys for collecting participant-recorded dietary data among Rwandan adults, demonstrating potential for assessing dietary quality for literate, higher income participants (Manners et al., 2025). However, for digitally marginalised respondents with incomplete primary education and low incomes, Manners et al. (2025) found percentage disagreement of consumed food groups between the text-message and weighed food record methods. In Kenya, another study collected participant-recorded anthropometric and dietary data for nomadic women and children using a picture-based smartphone application, yet the dietary data component was not validated (Jensen et al., 2023). The third study validated, with adolescent girls in Ghana, the use of a mobile artificial intelligence dietary assessment application using participant-recorded photos of foods consumed; however, it was a prospective method validated for quantified nutrient intakes in an urban context (Folson et al., 2023).

Interactive Voice Response (IVR) systems using mobile phones, have considerable potential for use in LMICs, enabling automated phone-based questionnaires where participants interact with high quality pre-recorded scripts by talking into the phone or pressing buttons on the phone’s keypad (Greenleaf et al., 2017; Tsoli et al., 2018). The advantages of the automated design of IVR are that it negates interviewer bias or error through consistent phrasing, pacing, and intonation of questions; it calls and records multiple participants simultaneously; it transcends low literacy levels; it can employ different languages; and process hundreds or thousands of calls in a single day (Lieberman & Naylor, 2012; Tsoli et al., 2018). These IVR features encompassing convenience, simplicity, confidentiality, and cost savings offer advantages compared to conventional enumerator-administered methods (Vecino-Ortiz et al., 2021). In high-income countries, IVR has proven effective for health monitoring, such as monitoring chronic pain (Lieberman & Naylor, 2012), and bowel symptoms (Lam et al., 2009). In LMICs, IVR has been utilised to elicit information on health practices in Bangladesh (Al Kibria et al., 2023), and household welfare indicators in Peru and Honduras (Ballivian et al., 2015).

The women’s dietary diversity score (WDDS) and minimum dietary diversity for women (MDD-W) are two standardised international indicators of dietary quality (FAO, 2021) validated as proxy indicators for micronutrient adequacy of women of reproductive age (WRA) in low-resource settings (Arimond et al., 2010; Verger et al., 2024). MDD-W is a key sustainable development goal indicator (FAO, 2025) and it, along with WDDS, have been in wide global use over the last decade for monitoring dietary changes, and assessing drivers and effects of nutrition policies, programmes and interventions on women’s dietary quality (O’Meara et al., 2025). To date, the validated versions of collecting women’s dietary diversity are enumerator-administered (Hanley-Cook et al., 2020). Although the Demographic and Health Surveys (DHS) have begun collecting women’s dietary diversity at scale, these resource-intensive surveys are only conducted every 4-10 years. To our knowledge, no validated participant-recorded IVR dietary data collection method currently exists for monitoring dietary quality of women in LMICs, especially amongst marginalised groups with low literacy.

This study aimed to: (i) refine a novel method of collecting women’s dietary diversity data using participant-recorded semi-quantitative list-based 24 h recalls administered via automated IVR on basic mobile phones with marginalised rural women in a sub-Sahara African context during the wet season, (ii) validate the IVR method, for assessing MDD-W, WDDS, and the percentage of women consuming unhealthy foods and beverages, using gold standard observed weighed food records (WFR) as the criterion method, and (iii) determine relative agreement compared with the conventional enumerator-administered 24-hour recall list-based method.

## Methods

### Study design

A cross-sectional study of women of reproductive age (18–49 years) with a biological child aged 12–23 months (n=156) was conducted during the wet season (August 29–November 3, 2022) in Apac and Kwania districts, Northern Region, Uganda. The study was nested within a larger project evaluating the use of mobile phones, wearable cameras, and accelerometers for collecting data on diets, energy expenditure, time use, and handwashing, and assessing the relationship between proximity to large waterbodies and women’s micronutrient adequacy. Reporting follows guidelines for inter-method accuracy (Cohen et al., 2016), and agreement studies (Kottner et al., 2011).

### Data collection pattern

Women’s food and beverage consumption was assessed using 15-h observed WFRs, enumerator-administered semi-quantitative 24-h list-based dietary recalls (24HR), and participant-administered semi-quantitative 24-h list-based dietary recalls via automated IVR. Data were collected over three consecutive days, using two possible patterns (**Figure 1**). Before data collection, a community sensitisation workshop was held. On day one, eligibility, anthropometric data, and a structured questionnaire were completed at a central village location; subsequent data were collected at participants’ homes/farms.

**Figure 1:**
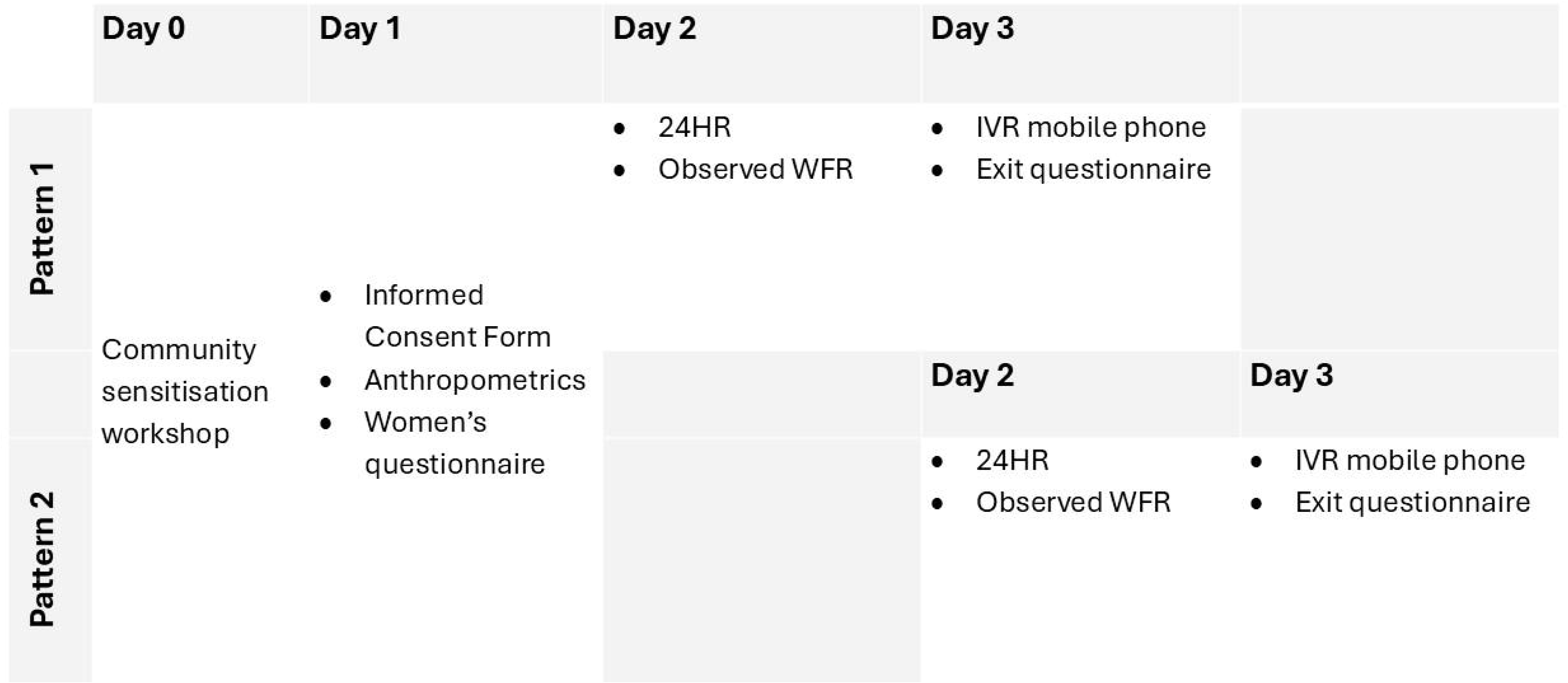
Data collection pattern. 24HR = semi-quantitative 24-h list-based dietary recall; WFR=weighed food record; IVR=Interactive Voice Response

On day two, an enumerator-administered 24HR was conducted in the morning, for the previous day’s intake, while the enumerator collected WFR data covering that day’s intake. On day three, the IVR recall was completed before other activities to capture dietary data for the same day as the WFR. A semi-structured exit questionnaire concluded the third day. For half the participants in each village, data collection began one day after the first half. Data were collected on a rolling basis across weekdays and weekends to account for day-of-week effects. For each participant, a different enumerator administered the WFR, IVR familiarisation, and exit questionnaire. All staff had tertiary qualifications and were trained beforehand.

### Participants and sampling

This study was conducted in Lango, Northern Uganda, a remote sub-region 7–9 h drive from Kampala, with a low human capital development index and high malnutrition rates (UBOS, 2016), reflecting significant disadvantage from historical conflict, unemployment, low education, and climate-related food insecurity. Apac and Kwania districts were purposively chosen for their proximity to large waterbodies (River Nile, Lake Kwania) supporting fisheries.

Sample size was calculated to detect a 10% difference in the mean probability of adequacy (MPA) in nutrient intakes, for women living ≤10 km and >10 km from large waterbodies (α=0.05, 80% power, SD=0.2) (Arimond et al., 2010), yielding 128 participants. This sample size was adjusted for clustering (design effect 1.03942) (Pries et al., 2019) and 10% attrition, resulting in a minimum sample size of 149. Due to enumerator team configurations, the target was set at 156. This sample size met the requirement of at least 100 participants to assess inter-method agreement using Bland-Altman plots (Bland & Altman, 1986). Mother-child dyads were selected via two-stage random sampling: 16 villages (eight near (<10km), eight far (>10km) from waterbodies) (O’Meara et al., 2021) were randomly chosen from 785 eligible villages based on lists compiled by local government officials, with three later excluded due to severe flooding (two close to waterbodies) (**Figure 2**). Villages located on floating islands or inside prisons or involved in formative research were also excluded. In each village, 12 eligible dyads (plus three reserves in selection order) were randomly selected from lists compiled by local leaders. Inclusion criteria included women aged 18–49 years, speaking Leblango, residing in the village, having no discernible disability that impaired participation in the study, available to participate during the week of data collection, and being the biological mother of a child aged 12-24 months. The inclusion criteria for the child were a singleton birth, had no discernible disability or illness affecting food consumption, currently eating solid foods, aged between 12 and 24 months and available during data collection. If insufficient dyads were found in a village, replacements dyads were randomly drawn from a nearby village. Sampling and random selection were calculated using R software ‘pwr’, ‘dplyr’, and ‘sample_n’ (R Core Team, 2024).

**Figure 2:**
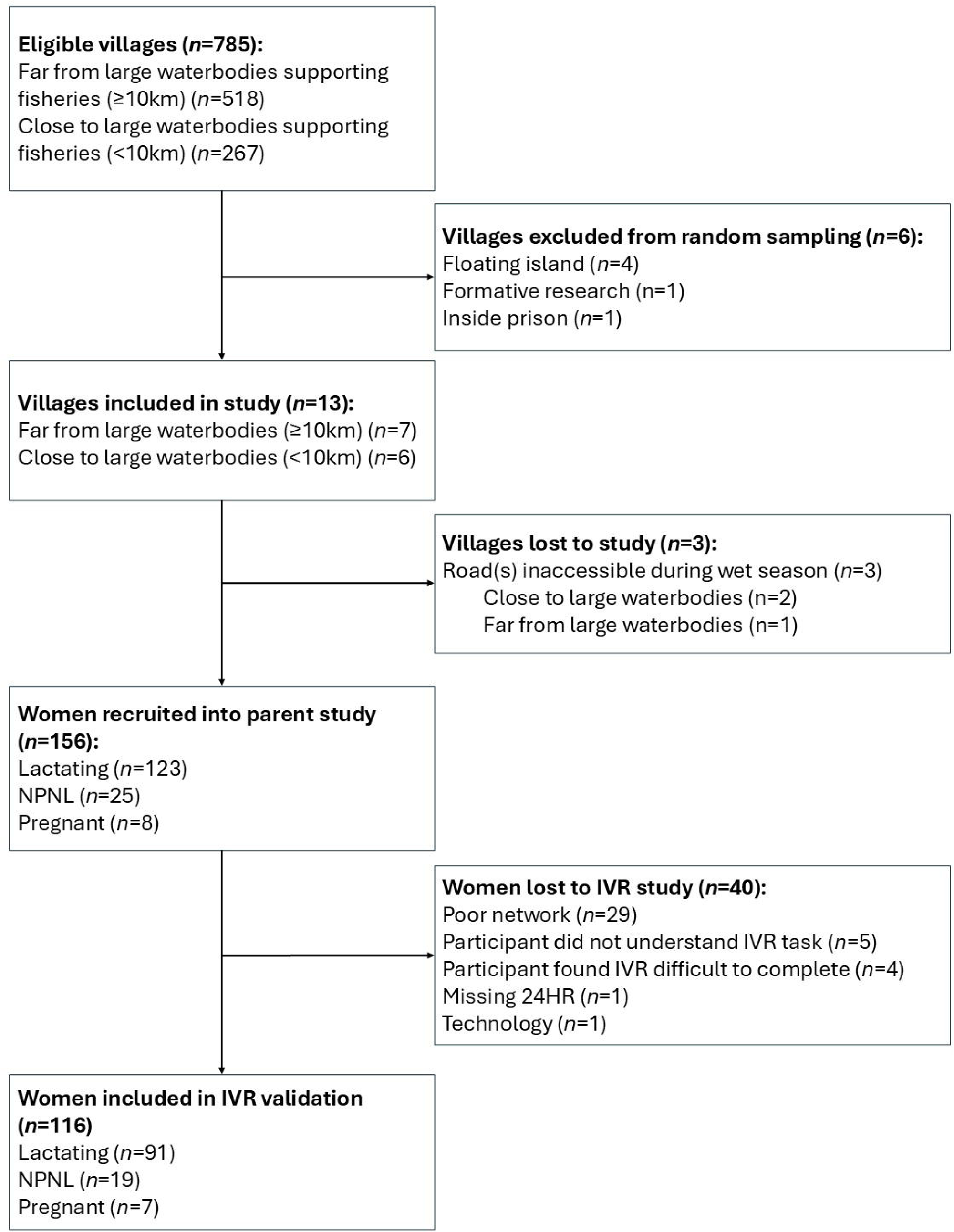
Flow diagram of study population. IVR=Interactive Voice Response; 24HR = 24-h list-based dietary recall: NPNL=non-pregnant/non-lactating

### Data collection tools and protocols Observed WFR and anthropometry

Each participant’s food and beverage intakes were estimated using a one-day WFR with dietary scales (±1g, Salter Disc Electronic Digital Scale Model 1036, UK) (Gibson, 2005). A trained female enumerator accompanied the participant from 06:30 to 21:00, weighing all foods and beverages served and left over. Recipe data were collected, including weighing raw ingredients, cooked food, and noting cooking methods. Foods or drinks consumed outside these hours were recorded using standard 24-h dietary recall methods using actual food, raw rice, or playdough models (Gibson and Ferguson, 2008). Duplicate anthropometric measurements (weight to 0.1kg, height to 0.5cm) were also taken (SECA weighing scales, Microtoise).

### Enumerator-administered questionnaires

Enumerators administered three structured questionnaires. The first covered socio-demographics, household wealth, food security, women’s empowerment, nutrition/health knowledge, and sources and biodiversity of local foods. The second was a semi-quantitative list-based 24-hour dietary recall for both participant and child, following Ugandan DHS-8 dietary diversity modules (ICF, 2023) and FAO (2021) guidelines. The exit questionnaire, using Likert and categorical scales, assessed participant perceptions of three of the four methods of dietary data collection used in the study, phone ownership/use, and willingness for future mobile-based studies, with open-ended questions for qualitative feedback. The first two questionnaires were collected using SurveyCTO software (Dobility Inc., v2022) on Android tablets, while the third was collected via paper-based methods.

### Novel automated Interactive Voice Response

The computerised IVR questionnaire format was adapted from the Uganda DHS dietary diversity modules for women and children (ICF, 2023), with identical questions of the IVR used for the semi-quantitative 24HR recall. IVR questions underwent contextualization (i.e., local food examples), translation into Leblango, audio recording by a female enumerator using a smartphone audio application, and pre-testing for clarity (**Supplementary Information [SI] Table 3**). The IVR included three list based semi-quantitative (yes/no) 24-h recall modules: child dietary diversity, women’s dietary diversity, and women’s handwashing practices. Audio files were uploaded to the engageSPARK platform (v2022), which was programmed with skip logic, invalid response handling, call retries and reconnects (engageSPARK, 2023). Project-issued basic phones (Techno T301) were used due to low phone ownership (**Figure 3**), and network-specific SIM cards were issued to address coverage issues. No airtime was needed since the IVR platform initiated calls.

**Figure 3:**
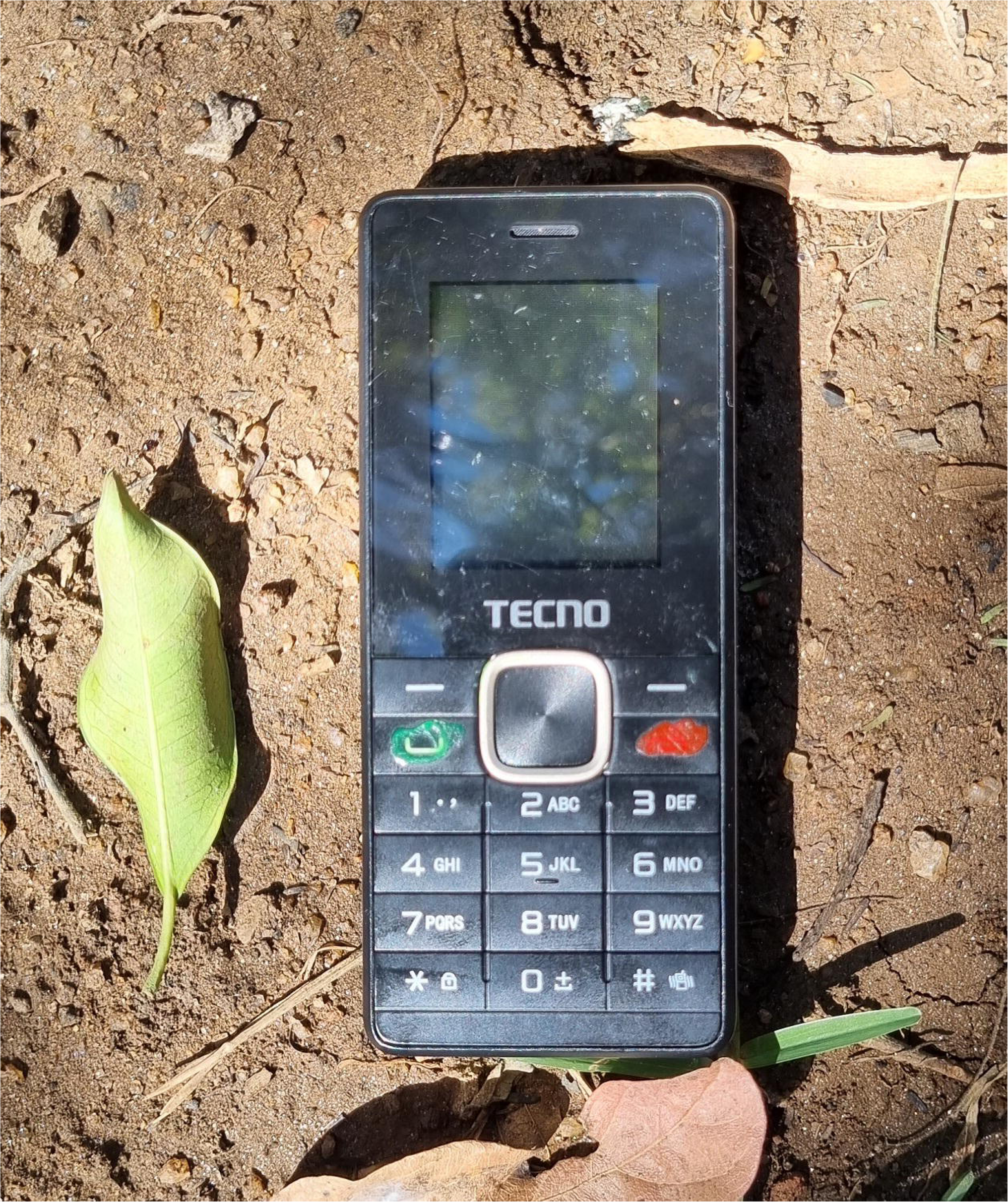
Basic push button mobile phone used in the study. Photo credit: O’Meara 2022

Formative research, including key informant interviews, focus groups and observational interactive pre-testing with local women, identified challenges such as low literacy, unfamiliarity with technology, and socio-cultural behaviours like gender-based violence around use of technology, as also reported in other projects in Northern Uganda (Mpiima et al., 2019). Lessons learnt from a prior stage of the parent project conducted in Eastern Uganda were also considered (Priebe et al., 2019). Protocols were adapted to include intensive community sensitisation, using a female voice in recordings, providing waist bags for discreet phone carrying, scheduling calls during late morning/midday when the woman was likely in the privacy of the farm, and using keypad responses.

The IVR protocol is outlined in **Figure 4**. An enumerator delivered and registered the phone in the morning, provided training, and triggered a practice call. The main IVR call was automatically placed 1 hour after registration with the platform, with repeated attempts every 15 min if needed. The IVR platform automatically guided the participant through the questions and would reconnect if disconnected due to poor network. Participants’ answered questions through keypad responses by pressing ‘1’ for yes, ‘3’ for no, and ‘8’ for ‘do not know’. The participant could choose to end the IVR at any time by hanging up and turning off the phone. If the participant failed to give an appropriate response after multiple attempts for the first two questions, then the system assumed that the participant did not understand the task and discontinued the call. Equipment was collected in the afternoon, and an exit survey administered. All IVR procedures followed digital ethical guidelines and privacy legislation.

**Figure 4:**
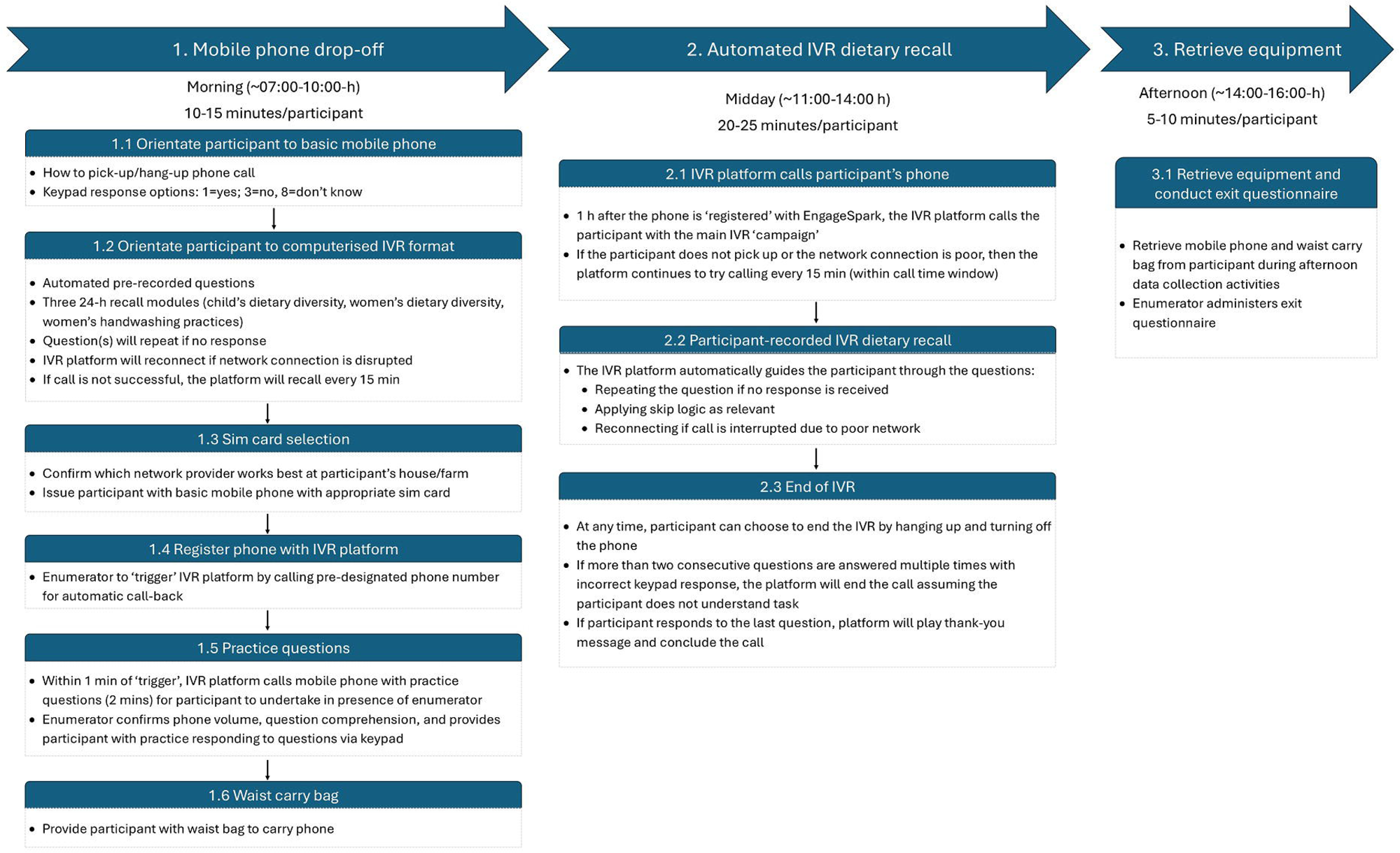
IVR data collection flowchart. IVR=Interactive Voice Response; min=minutes.

### Data processing

IVR response data were downloaded from engageSPARK website in Excel. Enumerator-administered questionnaires were cleaned on SurveyCTO before download; all other data were double-entered into Excel. Final cleaning was done in Excel and R. Dietary intake was coded into food groups, and WDDS was calculated for each method (WFR, 24HR, IVR) using the ten FAO-defined food groups (FAO, 2021). Discretionary foods from the Ugandan DHS-8 were coded as unhealthy foods and beverages (ICF, 2023). WFR data followed standard processing, including calculation of ingredient portion sizes from average household recipes (Gibson, 2005), with foods grouped by portion size cut-offs (≥1g/meal and ≥15g/meal). The ≥1g cut-off was used in addition to the conventional ≥15g (FAO, 2021) because the IVR asked women to report foods ‘no matter how small’ (**SI Table 3**). The percentage achieving MDDW (≥5 of 10 food groups) was calculated (FAO, 2021). Proportions living below $1.90/day (Schreiner, 2016), with severe food insecurity according to household food insecurity experience scores (FAO, 2013) or classified as thin (BMI <18.5) or overweight/obese (BMI ≥25) were also determined. Administrative data was used to determine time of IVR call, IVR reconnects, network connectivity, number of network providers per village, and IVR call duration.

### Statistical analysis

The primary outcomes were MDD-W, WDDS, and percentage of women consuming unhealthy foods and beverages. Cases with no data for IVR or 24HR were excluded. If a food was recorded on the WFR but the weight was missing, then substitute weights were input based on medians calculated from household recipes. For the IVR, completing up to the end of the women’s dietary 24-hour recall period was required for inclusion (question 65) (**SI Table 3**). Multiple statistical tests were used to triangulate validity as recommended (Lombard et al., 2015) and detailed below Wilcoxon signed-rank and McNemar’s chi-square tests compared IVR with WFR (same day) and 24HR (different day), for WDDS (ordinal) and MDDW (dichotomous), respectively (Bulungu et al., 2021; Hanley-Cook et al., 2020). Agreement between IVR and WFR was assessed using weighted Cohen’s kappa for WDDS and simple Cohen’s kappa for MDDW and food groups (Bulungu et al., 2021; De Bruyn et al., 2019; Hanley-Cook et al., 2020). Kappa values were interpreted as: ≤0 poor, 0.01–0.20 slight, 0.21–0.40 fair, 0.41–0.60 moderate, 0.61–0.80 substantial, 0.81–1.00 almost perfect (Landis & Koch, 1977). Because kappa values can be prevalence sensitive, resulting in the ‘kappa paradox’ of high percentage agreement but low kappa value, food groups with cell sizes <5 were excluded from kappa analysis. Food group/sub-group proportions were compared using McNemar’s test, and misreporting was assessed with 2×2 tables (Hanley-Cook et al., 2024). Statistical outputs were interpretated for acceptability to predict dietary intake according to Lombard et al. (2015). Socio-demographics of IVR completers versus non-completers were compared using Mann-Whitney (continuous) and Fisher Exact (categorical) tests (Bulungu et al., 2021).

WDDS agreement between IVR and WFR was evaluated with Bland-Altman plots and limits of agreement (LOA) (Bland and Altman, 1986; Bulungu et al., 2021; Hanley-Cook et al., 2024). A receiver operator characteristic analysis compared IVR’s ability to predict MDDW versus WFR (Hanley-Cook et al., 2024, 2020), with area under the curve (AUC) >0.70 indicating satisfactory accuracy (Akobeng, 2007). Analyses were conducted in R (v4.4.1) using ‘dplyr’, ‘tidyverse’, ‘psych’, ‘ggplot2’, ‘BlandAltmanLeh’, ‘pROC’, and ‘plotROC’ packages (R Core Team, 2024), with significance at p<0.05.

### Ethical considerations

Ethical approval was obtained from the University of Greenwich Faculty of Engineering and Science Ethics Committee (21.3.8.i.e), Uganda National Council for Science and Technology (A24ES), Clarke International University in Uganda (CIUREC/0066), and London School of Hygiene and Tropical Medicine (26645) IRB. Approval for digital data collection platforms (engageSPARK and SurveyCTO) was granted in line with UK data protection laws. Written prior informed consent or thumbprint was obtained from all participants. Modest compensation in kind was given to all participants for their time.

## Results

### Sample characteristics

A total of 156 women participated in the study. Of these, 74.4% (n=116) completed the IVR, while 25.6% (n=40) were lost to the study (**Figure 2**; **Table 1**). Losses included 18.6% (n=29) due to poor network, 5.8% (n=9) for user-related reasons (e.g., difficulty understanding or completing the task), 0.6% (n=1) due to technology (e.g., flat battery), and 0.6% (n=1) due to missing 24HR.

**Table 1:**
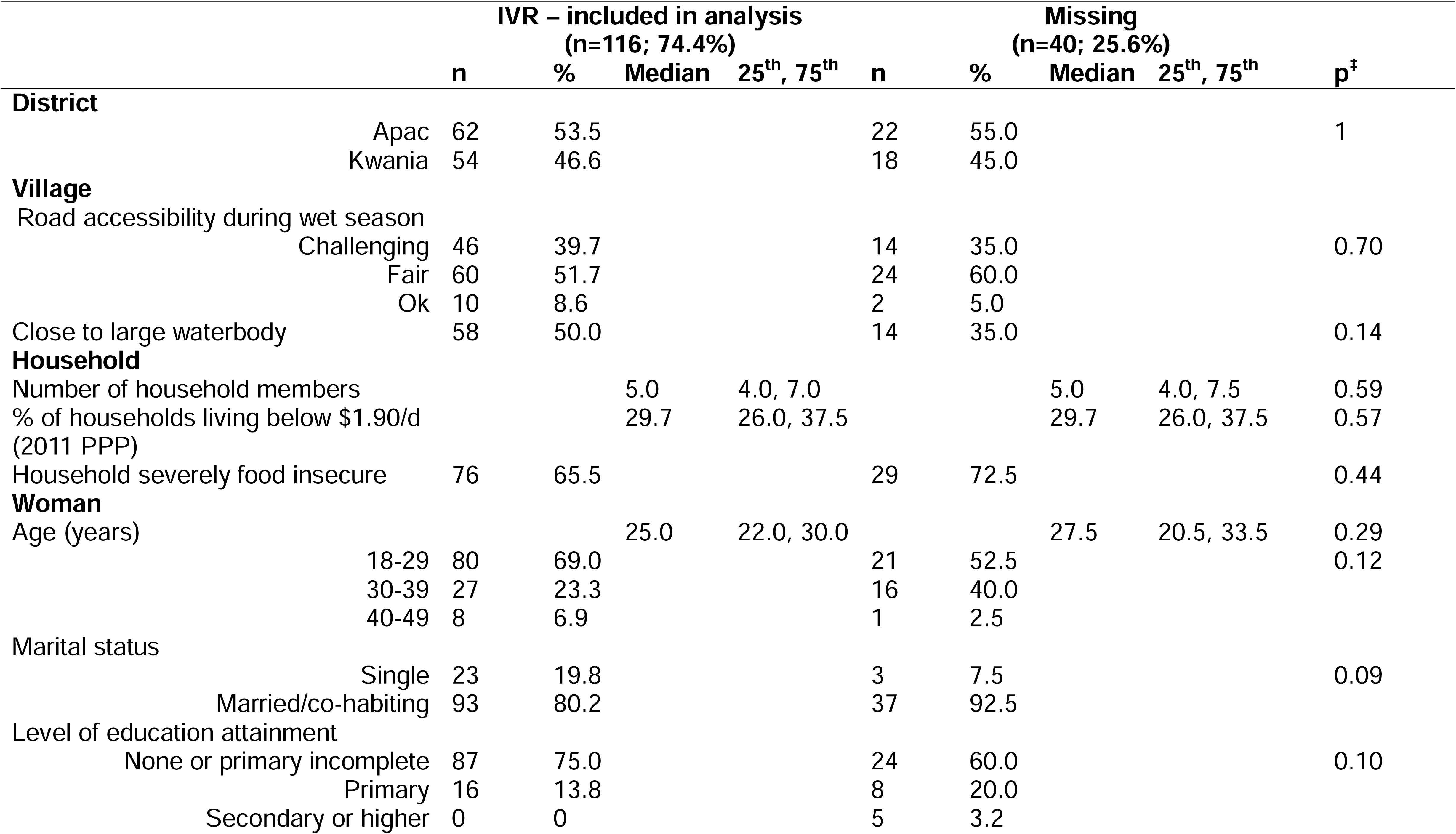
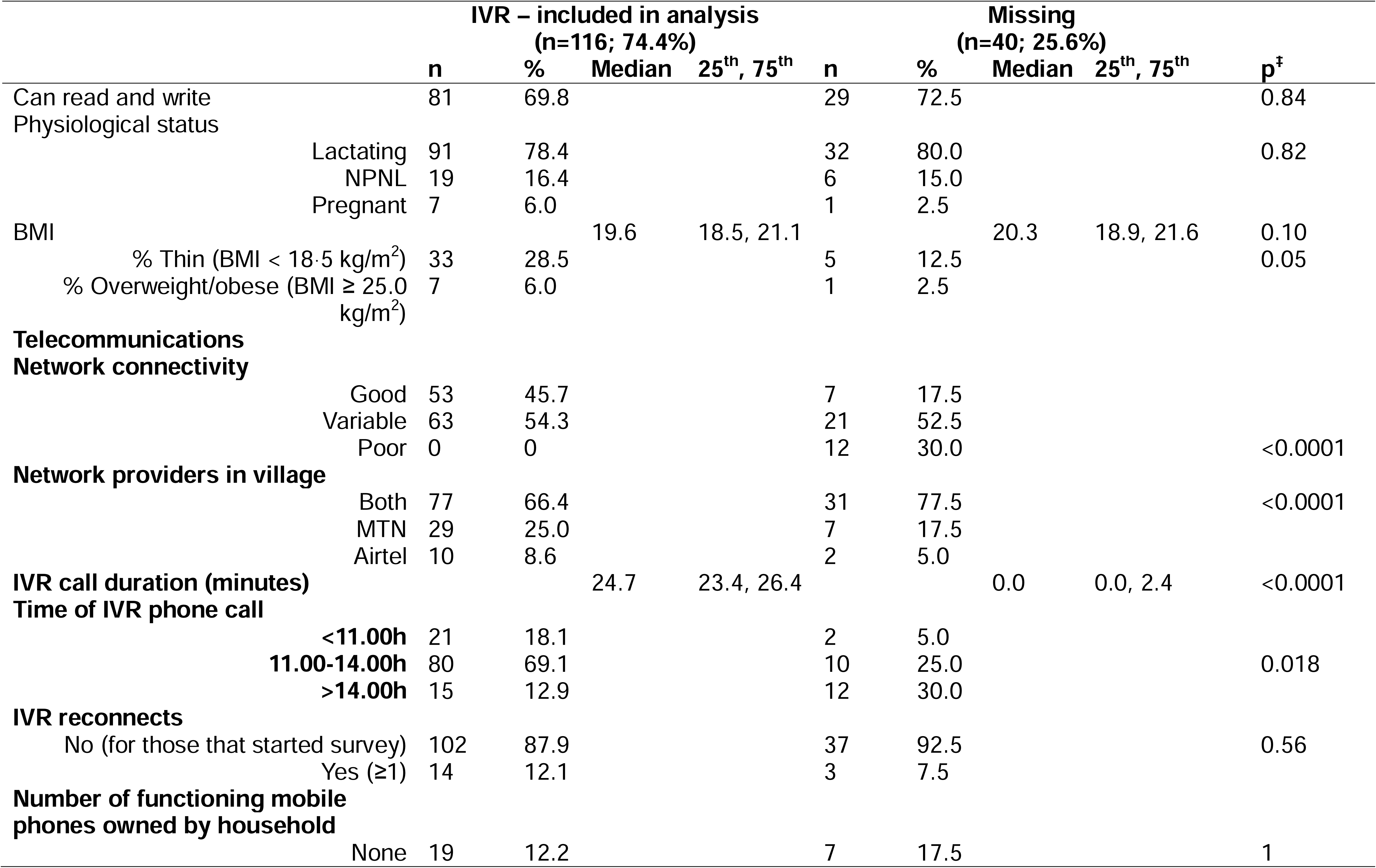
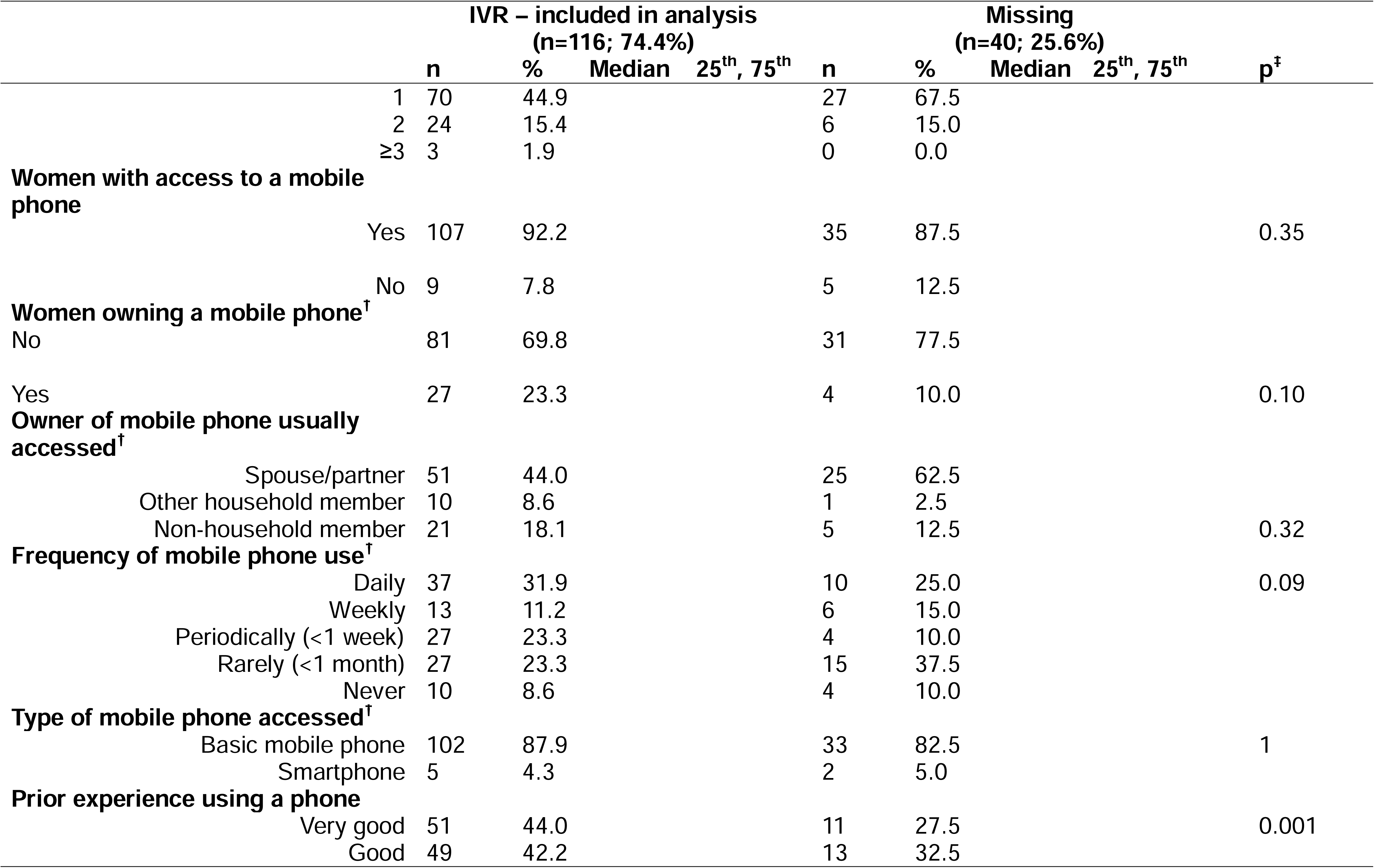
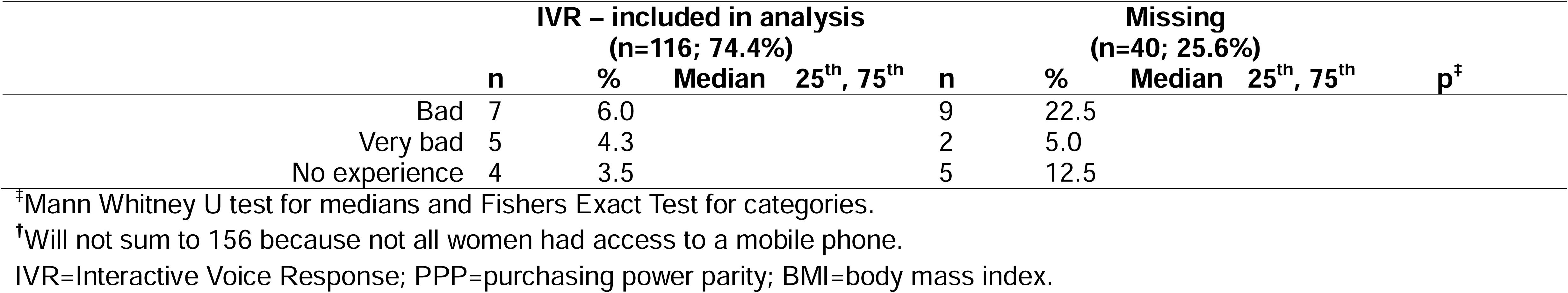
Mobile phone and IVR characteristics of all women in the study (n=156) and between those that were included in the analysis (n=116) and lost to the study (n=40).

Basic descriptive statistics (**Table 1**) showed that 53.8% of participants lived in Apac district, and 46.2% resided near large waterbodies, with 38.5% facing poor road access during the wet season. The average household size was five, and a median of 29.7% lived below $1.90 per day. Severe food insecurity affected 67.3% of households. The median age was 26 years; 83.3% were married, 78.8% were lactating, 71.2% had not completed primary school, and 29.5% were illiterate. Additionally, 24.4% were classified as thin. No statistically significant differences in these variables were found between women who completed the IVR and those lost to the study.

Telecommunications data showed 53.9% of women had variable and 7.7% poor network connectivity. Two network providers covered the villages of 69.2% of participants. The median IVR call lasted 24.3 minutes, with 57.7% completing the IVR between 11.00-14.00h. Among those who started and completed the IVR, 10.9% were reconnected at least once due to network dropout. In 83.3% of cases, the household owned a mobile phone. Although 71.8% of women did not own a phone, 91.0% had access to a phone. Of women with access to a phone, 86.5% had access to a basic mobile phone, and 55.8% had access toa phone owned by another household member. Mobile phone use was infrequent for 35.9% of women, and 20.6% had no or negative prior experience. Five telecommunication variables were statistically significant between IVR completers and non-completers: poor network connectivity, single network provider, shorter call duration, call time after 14.00h, and poor prior mobile phone experience were associated with lower completion.

### Inter-method agreement for WDDS

There were no significant differences in mean WDDS comparing the IVR estimates with those of the WFR (1g,15g; same day); however, it was significantly different between IVR and the 24HR (different day) measures (**Table 2**). Agreement between the IVR and the WFR was moderate for 1g and fair for 15g, according to weighted Cohen’s kappa. Although the Bland-Altman analysis demonstrated no difference, the LOA were wide, and the plots visualise a proportional bias with the two methods disagreeing more at lower (1g, 15g) and higher (1g) WDDS values (**Table 3**; **Figure 5**).

**Figure 5:**
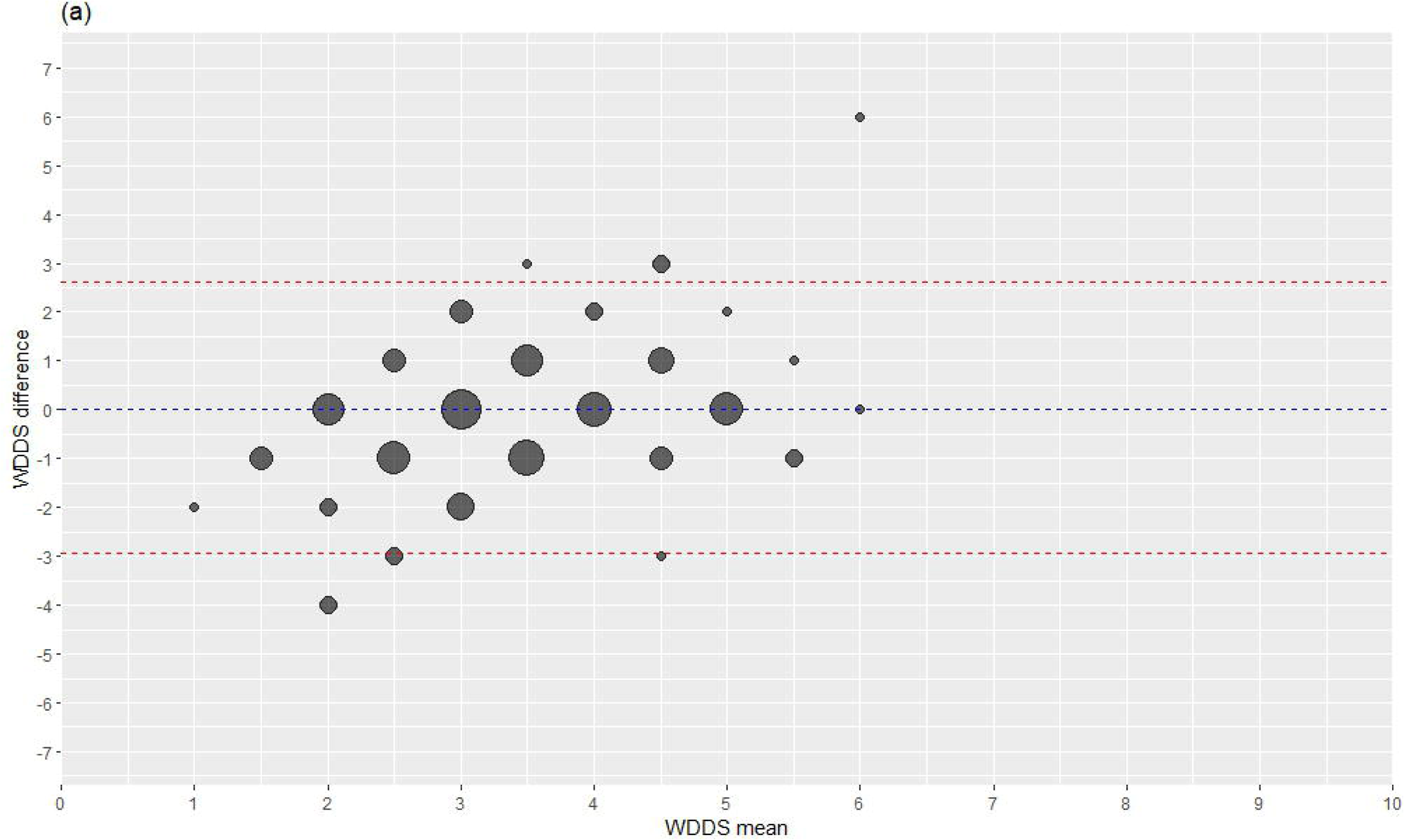
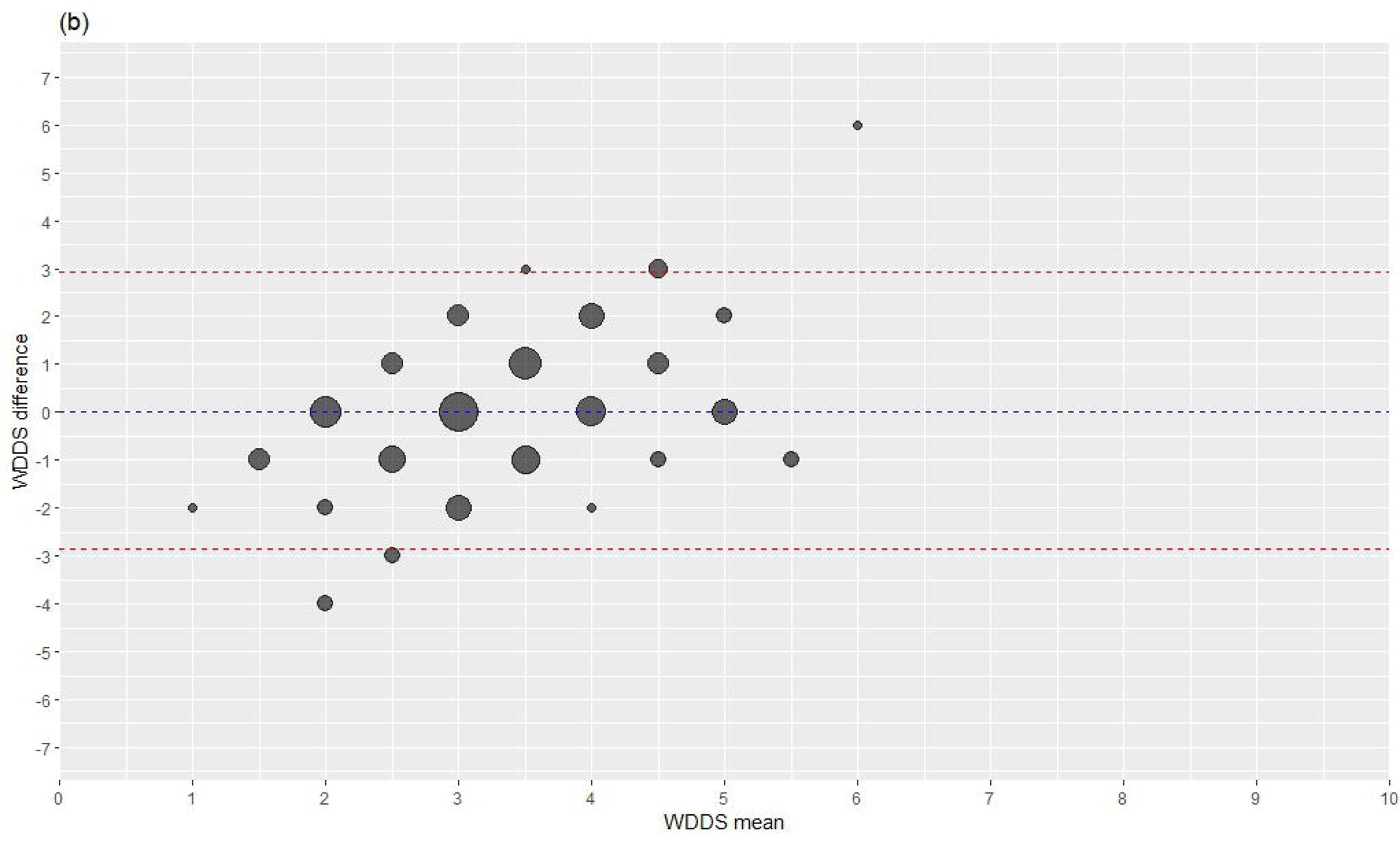
Bland Altman plot LOA between WDDS of WFR for (a) 1g and (b) 15g minimum requirements, and the IVR for participants (n=116). The size of the bubbles is proportional to the number of participants. LOA=limits of agreement; WDDS=women’s dietary diversity score; WFR=weighed food record; IVR=Interactive Voice Response

**Table 2:**
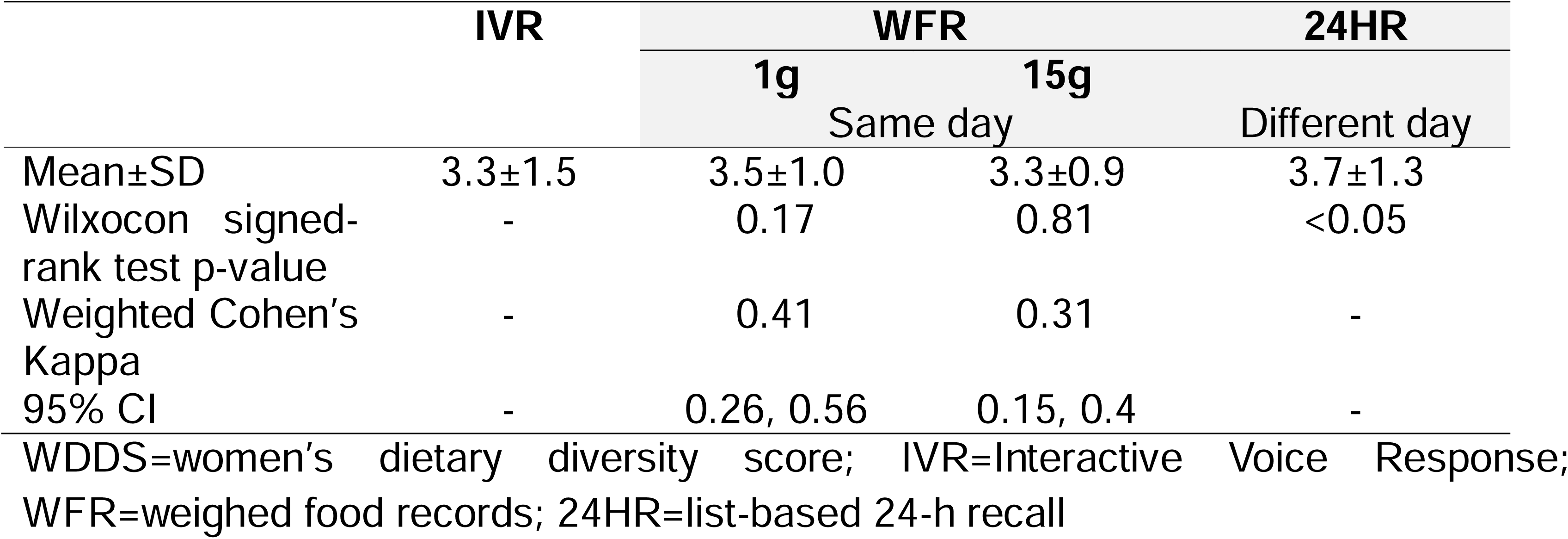
Descriptive statistics and statistical agreement of the WDDS for the participants (n=116) between the IVR compared with WFR and 24HR.

**Table 3:**
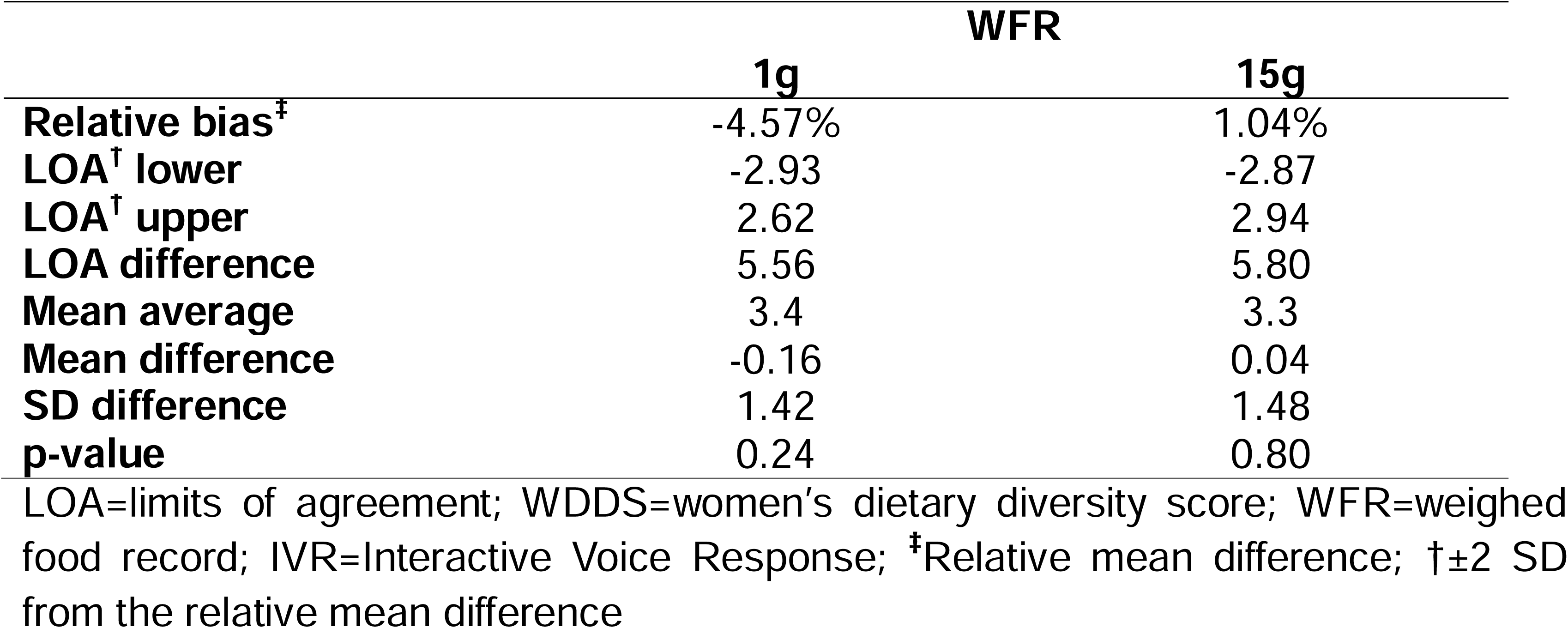
Inter-method comparisons of the relative bias and LOA (mean values and standard deviations) of the WDDS for the participants (n=116) between the WFR at 1g and 15g minimum requirements and the IVR, using Bland Altman analysis.

|  | WFR |  |
| --- | --- | --- |
|  | 1g | 15g |
| Relative bias<sup>‡</sup> | -4.57% | 1.04% |
| LOA<sup>†</sup> lower | -2.93 | -2.87 |
| LOA<sup>†</sup> upper | 2.62 | 2.94 |
| LOA difference | 5.56 | 5.80 |
| Mean average | 3.4 | 3.3 |
| Mean difference | -0.16 | 0.04 |
| SD difference | 1.42 | 1.48 |
| p-value | 0.24 | 0.80 |

LOA=limits of agreement; WDDS=women's dietary diversity score; WFR=weighed food record; IVR=Interactive Voice Response; <sup>‡</sup>Relative mean difference; <sup>†</sup>±2 SD from the relative mean difference

### Inter-method agreement for MDD-W

The percent of agreement between the IVR and WFR (1g, 15g) were high (>80%). According to simple Cohen’s kappa, agreement was moderate and fair for 1g and 15g, respectively (**Table 4**). The percentage of women achieving MDD-W was not statistically significant between IVR compared with WFR 1g or 24HR; however, the MDD-W was statistically significant between IVR compared with WFR 15g (**Table 5**). The AUC for ability of the IVR to predict MDD-W was satisfactory at 0.80 (WFR 1g) and 0.78 (WFR 15g) (**Figure 6**).

**Figure 6:**
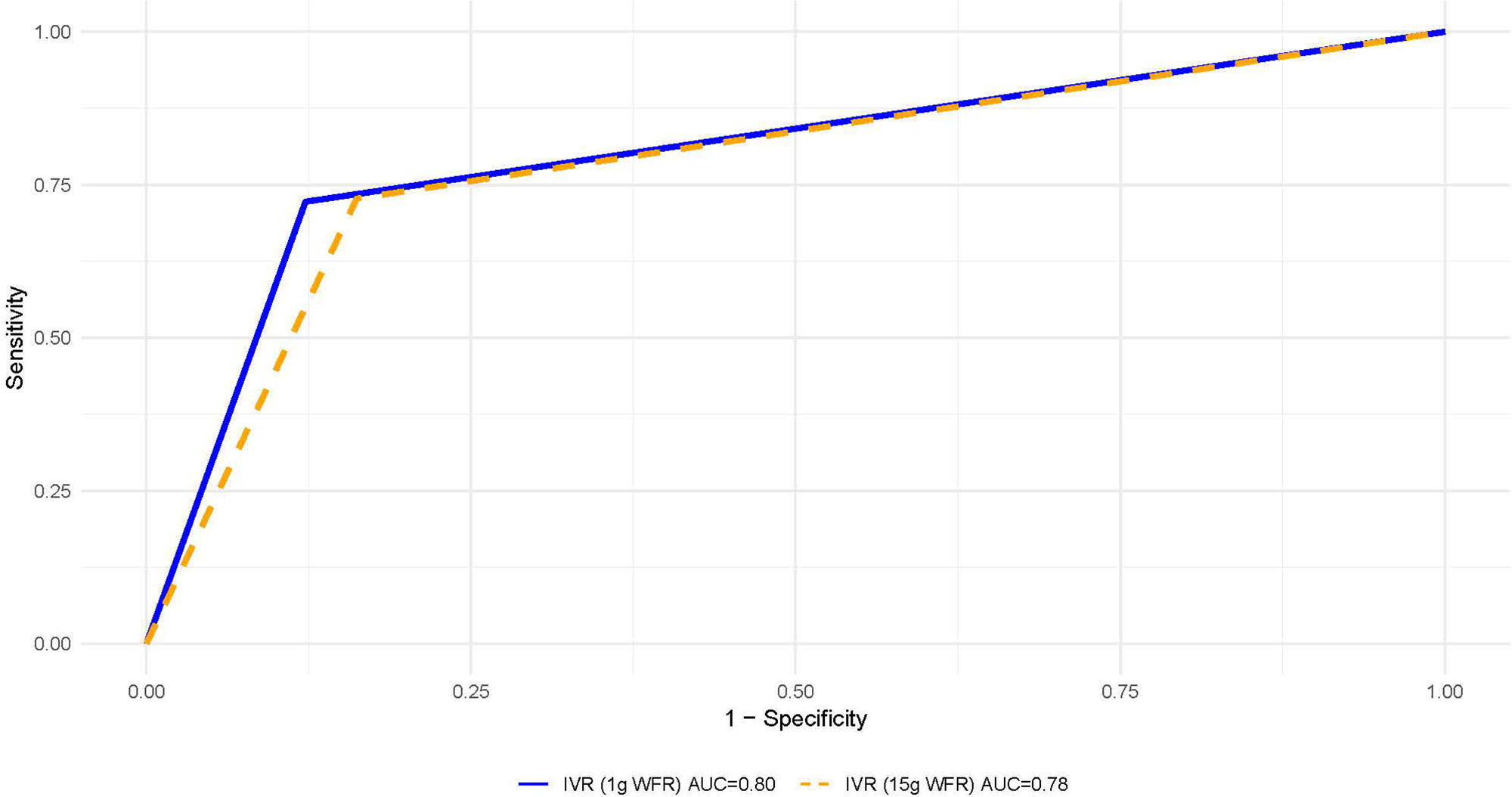
Receiver operating characteristic analysis for dichotomous MDD-W measured by IVR, as compared with WFR at 1g (AUC=0.80) and 15g (AUC=0.78) minimum requirement for participants (n=116). MDD-W=minimum dietary diversity for women; AUC=Area under the curve; IVR=Interactive Voice Response; WFR=weighed food records

**Table 4:**
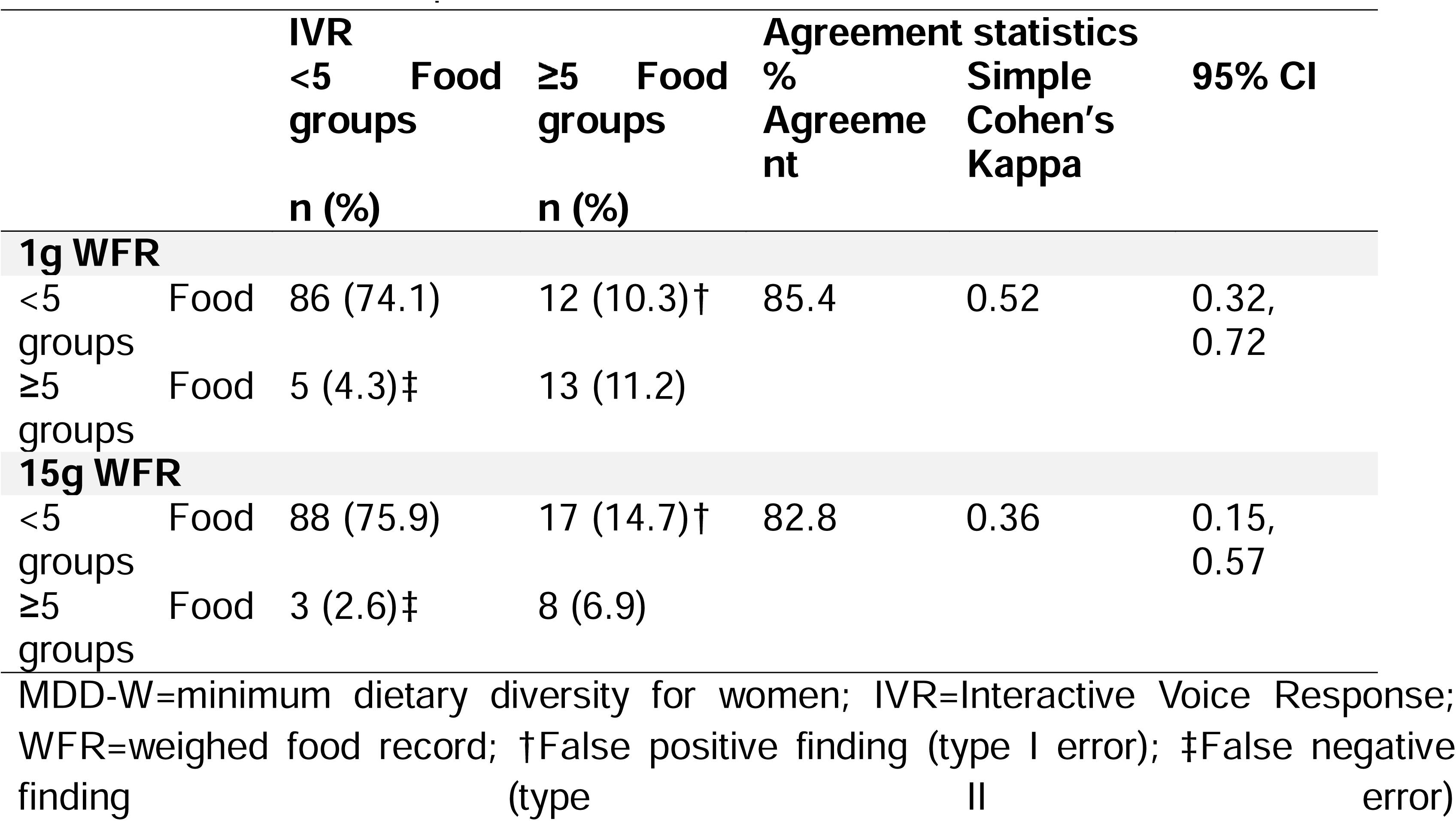
Agreement of the participant’s MDDW (n=116) between IVR and the WFR at 1g and 15g minimum requirements.

**Table 5:**
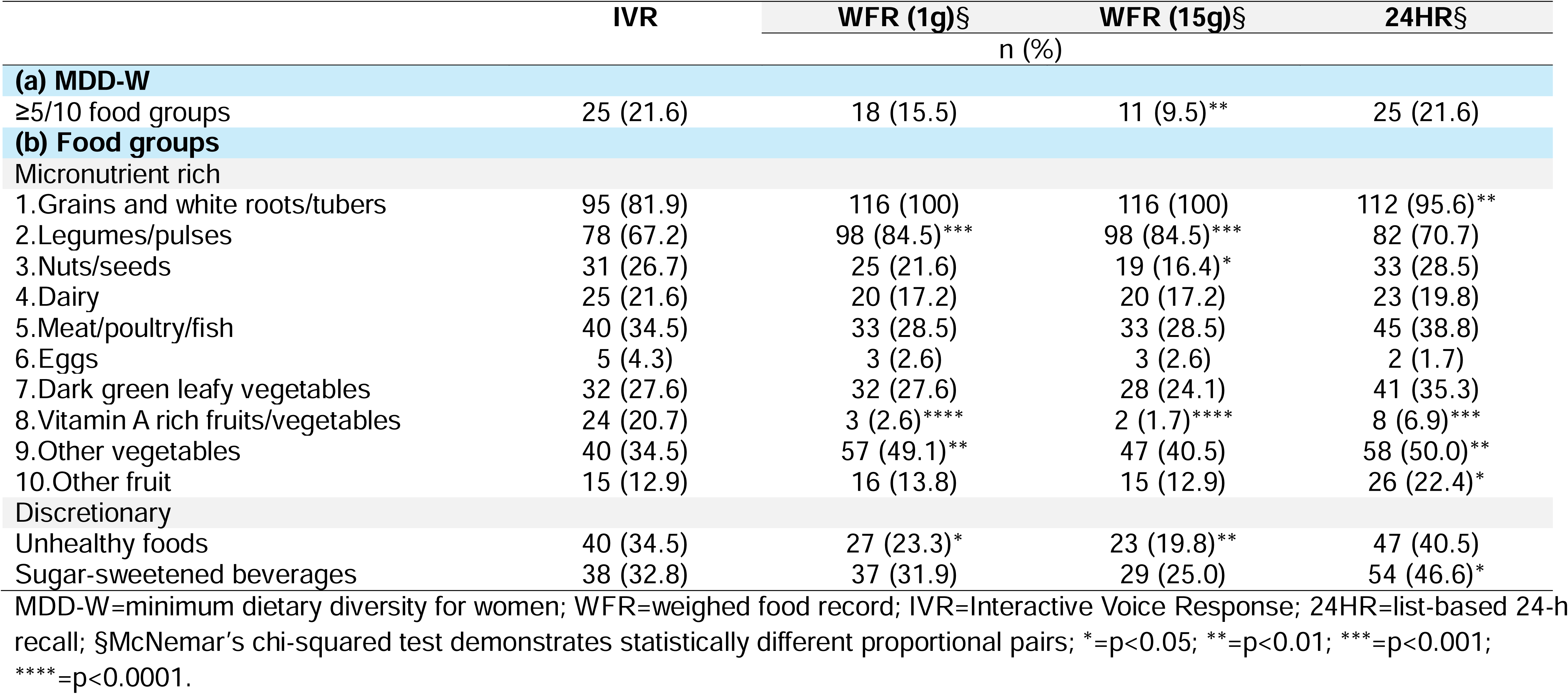
Percentage of women (n=116) having achieved MDD-W (a), and consumed food groups (b), of IVR compared with WFR (1g and 15g) and 24HR.

|  | IVR | WFR (1g)§ | WFR (15g)§ | 24HR§ |
| --- | --- | --- | --- | --- |
|  | n (%) |  |  |  |
| (a) MDD-W |  |  |  |  |
| ≥5/10 food groups | 25 (21.6) | 18 (15.5) | 11 (9.5)** | 25 (21.6) |
| (b) Food groups |  |  |  |  |
| Micronutrient rich |  |  |  |  |
| 1.Grains and white roots/tubers | 95 (81.9) | 116 (100) | 116 (100) | 112 (95.6)** |
| 2.Legumes/pulses | 78 (67.2) | 98 (84.5)*** | 98 (84.5)*** | 82 (70.7) |
| 3.Nuts/seeds | 31 (26.7) | 25 (21.6) | 19 (16.4)* | 33 (28.5) |
| 4.Dairy | 25 (21.6) | 20 (17.2) | 20 (17.2) | 23 (19.8) |
| 5.Meat/poultry/fish | 40 (34.5) | 33 (28.5) | 33 (28.5) | 45 (38.8) |
| 6.Eggs | 5 (4.3) | 3 (2.6) | 3 (2.6) | 2 (1.7) |
| 7.Dark green leafy vegetables | 32 (27.6) | 32 (27.6) | 28 (24.1) | 41 (35.3) |
| 8.Vitamin A rich fruits/vegetables | 24 (20.7) | 3 (2.6)**** | 2 (1.7)**** | 8 (6.9)*** |
| 9.Other vegetables | 40 (34.5) | 57 (49.1)** | 47 (40.5) | 58 (50.0)** |
| 10.Other fruit | 15 (12.9) | 16 (13.8) | 15 (12.9) | 26 (22.4)* |
| Discretionary |  |  |  |  |
| Unhealthy foods | 40 (34.5) | 27 (23.3)* | 23 (19.8)** | 47 (40.5) |
| Sugar-sweetened beverages | 38 (32.8) | 37 (31.9) | 29 (25.0) | 54 (46.6)* |

MDD-W=minimum dietary diversity for women; WFR=weighed food record; IVR=Interactive Voice Response; 24HR=list-based 24-h recall; §McNemar's chi-squared test demonstrates statistically different proportional pairs; \*=p<0.05; \*\*=p<0.01; \*\*\*=p<0.001; \*\*\*\*=p<0.0001.

### Inter-method agreement for food groups, and unhealthy foods and beverages

The proportion of women consuming unhealthy beverages was not statistically different between IVR compared with WFR (1g, 15g), but did differ from 24HR. Unhealthy food intake differed significantly between IVR and WFR but not 24HR (**Table 5; SI Table 1**). Agreement for unhealthy foods and beverages was 75.0% or higher, with moderate Cohen’s kappa values (**Table 6**). Food group agreement ranged from 73.3% to 96.6%, highest for fish/poultry/meat and dark green leafy vegetables (substantial), and lowest for other fruit and vitamin A-rich fruits/vegetables (poor and slight, respectively). Most false positives were for vitamin A-rich vegetables and sweets, whereas false negatives were more common for grains and roots/tubers (**SI Table 2**).

**Table 6:**
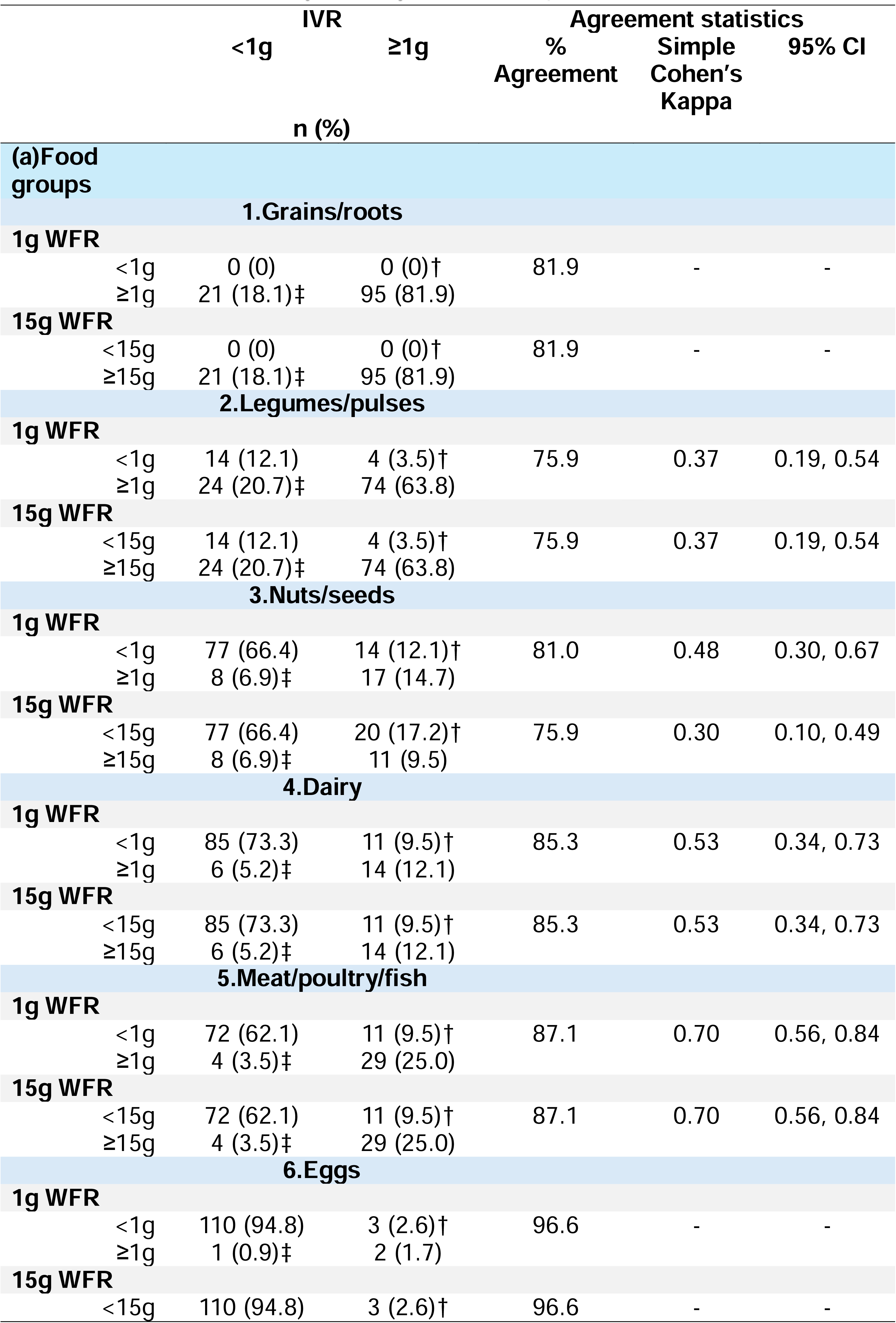
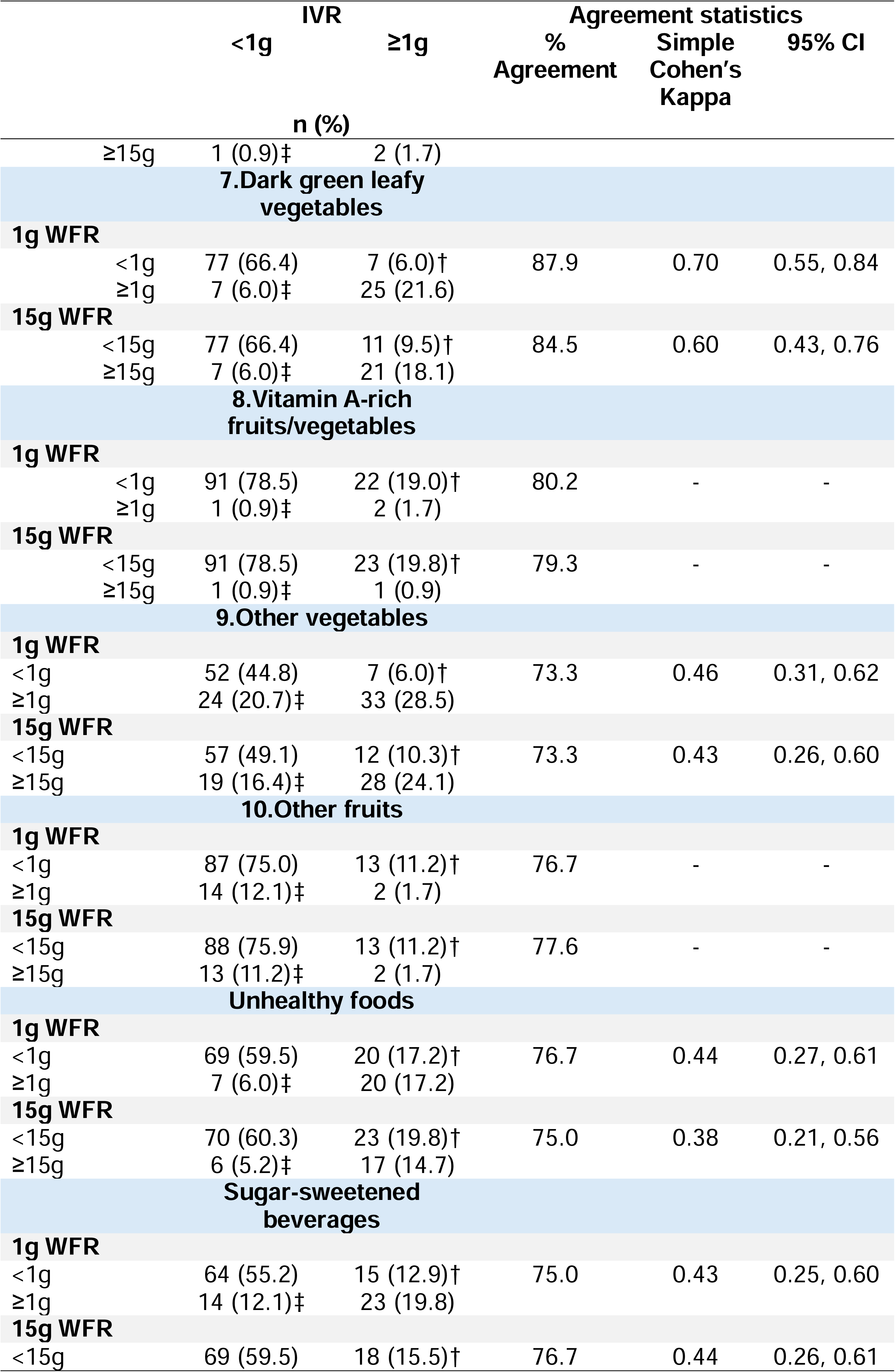
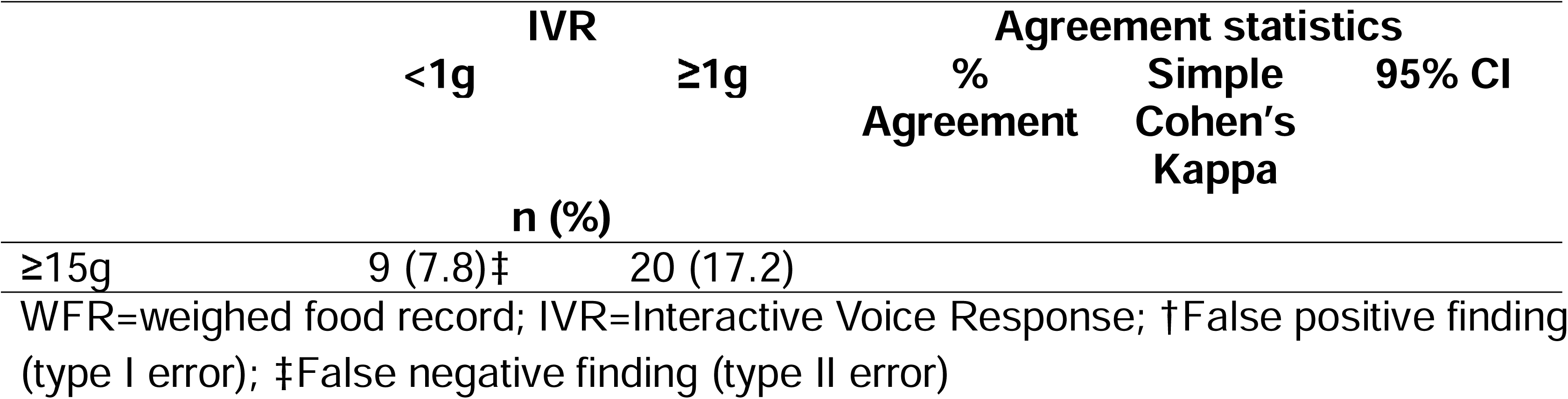
Inter-method agreement of food groups eaten by participants (n=116) between IVR and WFR at 1g and 15g minimum requirements.

### Acceptability

Most women (81.4%) reported they had a positive IVR experience, and 97.4% were willing to use IVR dietary reporting in future studies. Qualitative feedback was mostly positive, with some reporting that the IVR was “Easy to use and answer” (ID=100) and “Excited since [I] had never used a phone before” (ID=52). Some reported challenges, including fear due to limited phone knowledge (ID=87), lack of confidence answering questions (ID=70), and concerns about call length and repeated questions (ID=97).

## Discussion

This study found that a novel, automated participant-recorded 24-h list-based dietary recall via IVR on basic mobile phones is acceptably accurate for assessing population-level MDD-W, WDDS, and percentage consuming unhealthy foods and beverages among rural digitally marginalised women (19–49 years) during the wet season in Northern Uganda, using criteria specified by Lombard et al. (2015). Here we demonstrate the feasibility and validity of IVR for monitoring women’s dietary quality in a challenging context, including heavy rains during the wet season (i.e., interruptions to network connectivity), gender-sensitivity around the use of mobile phones, and low literacy and limited phone experience among rural women, supporting the robustness and usefulness of this method for collecting automated dietary data from marginalised population groups. Most women successfully completed the IVR, with completion associated with better network coverage, calling during morning/midday, longer call times, and prior positive mobile phone experience. The IVR estimated WDDS mean and proportion achieving MDD-W were not statistically different compared with the gold standard WFR, indicating the potential for this innovative automated mobile-phone based method to be used in place of conventional enumerator-administered methods for monitoring key international dietary quality indicators.

This is the first study to validate automated IVR dietary assessment using basic mobile phones among rural women in a sub-Saharan context, making direct comparisons difficult. We found a high percentage agreement of the IVR for predicting MDD-W against the WFR (84.4%; same day), similar to a study validating the ability of enumerator-administered mobile phone versus in-person surveys to predict MDD-W in Kenya (74.4%; different day) (Lamanna et al., 2019); however, our level of agreement may be slightly higher due to comparison with same day and WFR data. The ability of our IVR to predict MDD-W (kappa=0.52; AUC=0.80) and mean WDDS (weighted kappa=0.41) is also similar to a study validating the ability of enumerator-administered list-based 24 h dietary recalls to predict MDD-W (kappa=0.51; AUC=0.79) and mean WDDS (weighted kappa=0.47) against observed WFRs in Cambodia, Ethiopia, and Zambia (Hanley-Cook et al., 2020). We found that participants slightly over reported unhealthy food consumption, aligning with that found by Manners et al. (2025) in Rwanda, which may reflect aspirational responses. Although these studies are not directly comparable, these results indicate that our findings are at least consistent with other validation studies of MDD-W and WDDS in LMICs using enumerator-administered mobile phone surveys and semi-quantitative list-based 24HR.

Mixed results in food group reporting are common with enumerator-administered list-based recalls in LMICs due to memory lapses, social bias, telescoping, and the criterion of foods in the list to be asked (Assefa et al., 2022; De Bruyn et al., 2019; Hanley-Cook et al., 2024; Lamanna et al., 2019). In this study, the IVR accurately identified women consuming nutrient-rich foods like fish, poultry/meat, DGLVs, dairy, and nuts/seeds compared to WFRs (**SI Table 2**). However, the IVR over-reported vitamin A-rich vegetables and under-reported white roots/tubers and other vegetables versus WFR and 24HR. This misreporting likely stemmed from confusion about what qualifies as orange-fleshed sweet potato, a distinction that usually requires trained enumerators and probing (FAO, 2021). The order of IVR questions may have contributed, as vitamin A-rich vegetables were asked about before white roots/tubers (**SI Table 3**). Social bias is unlikely since nutrition knowledge on vitamin A-rich foods was low (<3.0%). Future IVR-administered recalls should carefully test question order and wording for these food groups. Like other studies, mixed dish ingredients, like tomatoes in sauces, were also sometimes missed, leading to under-reporting (Hanley-Cook et al., 2024).

While validation studies of mobile phone-based dietary tools in LMICs are limited, short recall periods (e.g., 24-hours versus 7-days) can capture out-of-period consumption and infrequently consumed foods may be missed or reported based on usual habits (Assefa et al., 2022; FAO, 2018). The IVR showed some over-reporting of infrequently consumed foods; however, results were comparable to 24HR, indicating similar performance compared with the conventional method. Although Bland-Altman analysis was not statistically significant, wide LOAs and visual plots indicate inter-method measurement errors were more common at lower and higher WDDS, limiting individual-level use (Gibson, 2005). However, this is not a significant drawback given that the MDD-W and WDDS indicators are only recommended for use at the population level (Arimond et al., 2010; FAO 2021).

By providing mobile phones and short training sessions, we were able to support the participation of some of the most marginalised women, negating non-coverage bias which can result in misidentification of population-level dietary quality rates by up to 25% in sub-Saharan African phone-based studies (Lamanna et al., 2019). Most participant loss was due to poor network connectivity, worsened by heavy rain, yet completion rates remained high (>70%), similar to IVR validation studies in HICs (Lam et al., 2009) and other phone-based surveys in sub-Saharan Africa (Manners et al., 2025; Lamanna et al., 2019). Tailoring SIM cards to the best local network and using automatic reconnect features improved completion, whilst providing waist bags helped women keep phones within hearing range during daily activities.

Individual training was important, especially for those unfamiliar with mobile phones. Women new to mobile phones were generally ‘excited’, though those with prior negative experiences using a mobile phone were less likely to complete the IVR. This indicates that some women will need extra support to enable equitable participation in future studies, especially in settings with high gender inequalities which privilege male participation (Al Kibria et al., 2023) or older women with higher asset ownership (Lamanna et al., 2019; Manners et al., 2025) when using phone-based methods.

Project phones were provided due to low mobile phone ownership among women. Although this necessitated an upfront investment, the cost of basic mobile phones was low (∼$10-15/phone) compared with smartphones or tablets (∼$150-200/tablet) routinely used in enumerator-administered methods (FAO, 2018). Moreover, basic mobile phones are often logistically easier to purchase in-country, and batteries are long-lasting. Many women had access to a phone via a household member, suggesting future studies may be able to use existing devices. As household and female mobile phone ownership rises, scalability will increase (GSMA, 2023).

Consistent with other mobile-based studies in LMICs (FAO, 2018; Folson et al., 2023; Jensen et al., 2023), we found high acceptability for IVR, with most women reporting a positive experience and willingness to use mobile phones for future dietary reporting. Prior positive experience with basic mobile phones was common and significantly associated with successful IVR completion. Smartphone ownership was rare, confirming the suitability of basic mobile phones in this low-resource context.

Careful contextualisation, translation, and simplification of IVR questions, plus community sensitisation and use of a female-voiced audio recording, were key for acceptability and safety (Mpiima et al., 2019), supporting the use of mobile phone-based dietary data collection in gender-sensitive settings. Phone-based methods were also less intrusive than other technologies like wearable cameras (Bulungu et al., 2023), supporting participant privacy.

This innovative digital IVR tool is well-suited for rapid high-frequency population-level monitoring of key international dietary quality indicators, and could be leveraged to fill dietary data gaps (Manners et al., 2025; Al Kibria et al., 2023; FAO, 2018; Vecino-Ortiz et al., 2021), track progress on global nutrition and food system goals (Fanzo et al., 2021; FAO, 2025; Schneider et al., 2023), and inform food policies for marginalised population groups in resource-scarce settings (O’Meara et al., 2025). The strength of IVR to provide real-time data (Ballivian et al., 2017; Al Kibria et al., 2023; Vecino-Ortiz et al., 2021) is particularly useful for policymakers developing timely and targeted policy responses during the first 1,000 days of life when nutrition impacts can be lifelong (Victora et al., 2008).

The nutrition risks of climate change, conflict and pandemics/epidemics are intensifying; however, funding for broad-scale food security, nutrition, and health monitoring is diminishing globally (FAO/IFAD/UNICEF/WFP/WHO, 2024). Therefore, the exponential growth in mobile phone network coverage, and mobile phone ownership in LMICs, including sub-Saharan Africa, will likely increase the future feasibility of the IVR method (GSMA, 2023; Greenleaf et al., 2017), increasing the cost-effectiveness of automated participant-recorded approaches (Manners et al., 2025; Al Kibria et al., 2023; Vecino-Ortiz et al., 2021).

Monitoring women’s dietary diversity can also serve as an early warning for food insecurity (IFPRI, 2002), especially in gender-unequal, low-resource settings where food is often privileged to other household members over young women (O’Meara et al., 2025). With prior recruitment and training, the ability of IVR to transcend low literacy and geographical barriers makes it ideal for monitoring dietary quality during times when physical access is constrained but food security and dietary quality is most at risk such as during wet ‘hungry’ seasons, population displacement, nomadic movement, severe weather events, conflicts, or disease outbreaks. Such high-frequency data could also track seasonal and environmental impacts on dietary quality, addressing the chronic scarcity of longitudinal data for marginalised population groups in LMICs (Meenakshi & Quisumbing, 2025), particularly in climate- and conflict-affected regions (O’Meara et al., 2025).

This study had several limitations. It is cross-sectional, and some villages were excluded because of flooding. Due to unforeseen resource constraints, repeated measures were not possible, limiting assessment of intra-individual reliability; however, prioritising sample size over repeated measures is recommended when estimating a population mean (Gibson, 2005). Like all dietary recall methods, self-reported approaches have inherent weaknesses, although the IVR performed comparably to the conventional method. The sample size was moderate, and the study was nested within a broader research project, which may have influenced participation and reporting.

Strengths of this study included the use of WFRs as the criterion method (Gibson, 2005), and multiple statistical tests to triangulate validity (Lombard et al., 2015). Extensive formative research informed protocol refinement, and the tool was validated during the wet season with marginalised rural women in a patriarchal context, demonstrating robustness under challenging conditions. The sample included a diverse mix of fisheries- and agro-pastoralist-dependent households and all reproductive physiological stages (lactating, pregnant, NPNL), supporting applicability for WRA across different rural livelihoods.

Future research needs include validation of the IVR tool for diverse contexts, other vulnerable groups such as adolescent girls, and for remote longitudinal monitoring. In the absence of a true ‘gold standard’ for dietary assessment validation studies, future research would also benefit from triangulating results with repeated measures and against a biomarker such as doubly labelled water (Lombard et al., 2015).

## Conclusion

This is the first study to validate a participant-recorded 24-h list-based dietary recall using IVR on basic mobile phones among women with low literacy and limited mobile phone ownership and experience during the wet season in sub-Saharan Africa. Compared to gold-standard WFRs and the in-person enumerator-administered 24HR, the IVR tool demonstrated valid population-level measurement of key international dietary quality indicators, with accuracy comparable to conventional methods. In this study we demonstrate that with short participant training, IVR’s voice-based, keypad-response format bypassed low literacy barriers and is feasible in resource-limited settings. This tool may be used for assessment of population dietary risk, trends over time, and group differences, offering a simple, automated solution for collecting dietary data from low literate, rural groups. As the risks of climate change, conflict, and infectious disease outbreaks intensify, such digital tools can support rapid, equitable monitoring and timely food security and nutrition policy responses, reducing reliance on costly in-person surveys. This approach could transform dietary surveillance for marginalised women who are often most at risk of malnutrition. Methodological refinements could further reduce misreporting. Overall, IVR is suitable for population-level monitoring of women’s dietary quality, but further validation is needed for different contexts, remote longitudinal use, and other vulnerable groups. Based on key international dietary quality indicators and low-cost basic mobile phones, with appropriate adaptation, this method can be generalisable to other LMICs.

## Data sharing

Data is available at Harvard Dataverse: “Replication Data for: validation of Interactive Voice Response (IVR) for collecting dietary data among low-literate women in Northern Uganda 2022”. R script for all analysis is available on github: https://github.com/l-c-omeara/ivr-validation.git.

## Acknowledgements

The authors would like to thank the women who participated in this study. We would also like to thank Elvis Mugabi of the Ichuli Institute for IVR programming assistance; Christine Hortinela and Avner Mizrahi of engageSPARK for IVR training and support; Stephen Young (NRI, University of Greenwich) for statistical guidance; Lora Forsythe (NRI, University of Greenwich) and Esther Nyapendi (Women of Uganda Network) for guidance on gender-sensitive use of mobile phones; and Julia de Bruyn (NRI, University of Greenwich) on validation of innovative tools in low-literacy groups.

## Author contributions

Conceptualization=KW, EF, JN, LO; Data curation=LO; Formal analysis=LO; Funding acquisition=KW, EF, JN; Investigation=LO; Methodology=LO, EF, KW, JN, PO; Project administration=LO, KW, JN; Resources=LO, KW, JN, PO; Software=LO; Supervision=KW, EF, PDS; Validation=LO, EF; Visualization=LO; Writing – original draft=LO; Writing – review and editing=LO, EF, KW, JN, PDS, PO.

## Declaration of use of generative AI

During the preparation of this work the main author (LO) used ChatGPT to refine R code, and Perplexity for grammar, spelling, and word count reduction. After using these tools, all authors reviewed and edited the content as needed and take full responsibility for the content of the published article.

