## Supplementary Information for "A novel automated participant-recorded dietary data collection method using low-cost mobile phones and Interactive Voice Response (IVR) with low-literacy women: a validation study in rural Uganda"

O'Meara et al

- **SI Table 1**: Percentage of women consuming food subgroups between data collection methods
- **SI Table 2**: Inter-method agreement of food sub-groups between IVR and WFR
- **SI Table 3**: IVR questionnaire, including skip logic and translation

**SI Table 1**: Percentage of women (*n*=116) consuming food sub-groups of IVR compared with WFR (1g and 15g) and 24HR.

| | IVR | WFR (1g)§ | WFR (15g)§ | 24HR§ |
|---|---|---|---|---|
| | | *n (%)* | | |
| **Food sub-groups** | | | | |
| Grains/roots | | | | |
| Grains | 71 (61.2) | 83 (71.6) | 83 (71.6) | 90 (77.6)** |
| Roots/tubers (white) | 45 (38.8) | 82 (70.7)**** | 82 (70.7)**** | 71 (61.2)**** |
| Meat/poultry/fish | | | | |
| Meat/poultry | 12 (10.3) | 9 (7.8) | 8 (6.9) | 11 (9.5) |
| Organs | 3 (2.6) | 3 (2.6) | 3 (2.6) | 0 (0) |
| Fish | 27 (23.3) | 26 (22.4) | 26 (22.4) | 35 (30.2) |
| Small protein (insects, fish eggs) | 8 (6.9) | 0 (0) | 0 (0) | 4 (3.5) |
| Vitamin A-rich fruits/vegetables | | | | |
| Vitamin A-rich vegetables | 20 (17.2) | 0 (0) | 0 (0) | 4 (3.5)*** |
| Vitamin A-rich fruits | 7 (6.0) | 3 (2.6) | 2 (1.7) | 4 (3.5) |
| Unhealthy foods | | | | |
| Fried foods | 27 (23.3) | 23 (19.8) | 22 (19.0) | 40 (34.5) |
| Sweets | 23 (19.8) | 5 (4.3)**** | 1 (0.9)**** | 15 (12.9) |
| Processed meat | 1 (0.9) | 0 (0) | 0 (0) | 1 (0.9) |
| Sugar-sweetened beverages | | | | |
| Soda | 8 (6.9) | 1 (0.9)* | 1 (0.9) | 16 (13.8) |
| Sugar with tea | 30 (25.9) | 36 (31.0) | 28 (24.1) | 38 (32.8) |
| Juice | 4 (3.5) | 0 (0) | 0 (0) | 3 (2.6) |

*MDD-W=minimum dietary diversity for women; WFR=weighed food record; IVR=Interactive Voice Response; 24HR=list-based 24-h recall; §McNemar's chi-squared test demonstrates statistically different proportional pairs; *=p<0.05; **=p<0.01; ***=p<0.001; ****=p<0.0001.*

**SI Table 2**: Inter-method agreement of food sub-groups eaten by participants (*n*=116) between IVR and WFR at 1g and 15g minimum requirements.

| | IVR | | Agreement statistics | | |
|---|---|---|---|---|---|
| | **<1g** | **≥1g** | **% Agree-ment** | **Simple Cohen's Kappa** | **95% CI** |
| | n (%) | | | | |
| **Food sub-groups** | | | | | |
| **Grains** | | | | | |
| **1g WFR** | | | | | |
| <1g | 18 (15.5) | 15 (12.9)† | 63.8 | 0.20 | 0.02, 0.38 |
| ≥1g | 27 (23.3)‡ | 56 (48.3) | | | |
| **15g WFR** | | | | | |
| <15g | 18 (15.5) | 15 (12.9)† | 63.8 | 0.20 | 0.02, 0.38 |
| ≥15g | 27 (23.3)‡ | 56 (48.3) | | | |
| **Roots/tubers (white)** | | | | | |
| **1g WFR** | | | | | |
| <1g | 33 (28.5) | 1 (0.9)† | 66.4 | 0.38 | 0.26, 0.51 |
| ≥1g | 38 (32.8)‡ | 44 (37.9) | | | |
| **15g WFR** | | | | | |
| <15g | 33 (28.5) | 1 (0.9)† | 66.4 | 0.38 | 0.26, 0.51 |
| ≥15g | 38 (32.8)‡ | 44 (37.9) | | | |
| **Meat/poultry** | | | | | |
| **1g WFR** | | | | | |
| <1g | 103 (88.8) | 4 (3.5)† | 95.7 | 0.74 | 0.52, 0.96 |
| ≥1g | 1 (0.9)‡ | 8 (6.9) | | | |
| **15g WFR** | | | | | |
| <15g | 103 (88.8) | 5 (4.3)† | 94.8 | 0.67 | 0.43, 0.92 |
| ≥15g | 1 (0.9)‡ | 7 (6.0) | | | |
| **Organs** | | | | | |
| **1g WFR** | | | | | |
| <1g | 110 (94.8) | 3 (2.6)† | 94.8 | - | - |
| ≥1g | 3 (2.6)‡ | 0 (0.0) | | | |
| **15g WFR** | | | | | |
| <15g | 110 (94.8) | 3 (2.6)† | 94.8 | - | - |
| ≥15g | 3 (2.6)‡ | 0 (0.0) | | | |
| **Fish** | | | | | |
| **1g WFR** | | | | | |
| <1g | 83 (71.6) | 7 (6.0)† | 88.8 | 0.68 | 0.52, 0.84 |
| ≥1g | 6 (5.2)‡ | 20 (17.2) | | | |
| **15g WFR** | | | | | |
| <15g | 83 (71.6) | 7 (6.0)† | 88.8 | 0.68 | 0.52, 0.84 |
| ≥15g | 6 (5.2)‡ | 20 (17.2) | | | |
| **Small protein** | | | | | |
| **1g WFR** | | | | | |
| <1g | 108 (93.1) | 8 (6.9)† | 93.1 | - | - |
| ≥1g | 0 (0)‡ | 0 (0) | | | |
| **15g WFR** | | | | | |
| <15g | 108 (93.1) | 8 (6.9)† | 93.1 | - | - |
| ≥15g | 0 (0)‡ | 0 (0) | | | |
| **Vitamin A-rich vegetables** | | | | | |
| **1g WFR** | | | | | |
| <1g | 96 (82.8) | 20 (17.2)† | 82.8 | - | - |
| ≥1g | 0 (0)‡ | 0 (0) | | | |
| **15g WFR** | | | | | |
| <15g | 96 (82.8) | 20 (17.2)† | 82.8 | - | - |

| | | | | | |
|---|---|---|---|---|---|
| ≥15g | 0 (0)‡ | 0 (0) | | | |
| **Vitamin-A rich fruits** | | | | | |
| **1g WFR** | | | | | |
| <1g | 108 (93.1) | 5 (4.3)† | 94.8 | - | - |
| ≥1g | 1 (0.9)‡ | 2 (1.7) | | | |
| **15g WFR** | | | | | |
| <15g | 108 (93.1) | 6 (5.2)† | 94.0 | - | - |
| ≥15g | 1 (0.9)‡ | 1 (0.9) | | | |
| **Fried foods** | | | | | |
| **1g WFR** | | | | | |
| <1g | 79 (68.1) | 14 (12.1)† | 79.3 | 0.39 | 0.19, 0.59 |
| ≥1g | 10 (8.6)‡ | 13 (11.2) | | | |
| **15g WFR** | | | | | |
| <15g | 80 (68.9) | 14 (12.1)† | 80.2 | 0.41 | 0.21, 0.61 |
| ≥15g | 9 (7.8)‡ | 13 (11.2) | | | |
| **Sweets** | | | | | |
| **1g WFR** | | | | | |
| <1g | 93 (80.2) | 18 (15.5)† | 84.5 | - | - |
| ≥1g | 0 (0.0)‡ | 5 (4.3) | | | |
| **15g WFR** | | | | | |
| <15g | 93 (80.2) | 22 (19.0)† | 81.0 | - | - |
| ≥15g | 0 (0.0)‡ | 1 (0.9) | | | |
| **Processed meat** | | | | | |
| **1g WFR** | | | | | |
| <1g | 115 (99.1) | 1 (0.9)† | 99.1 | - | - |
| ≥1g | 0 (0)‡ | 0 (0) | | | |
| **15g WFR** | | | | | |
| <15g | 115 (99.1) | 1 (0.9)† | 99.1 | - | - |
| ≥15g | 0 (0)‡ | 0 (0) | | | |
| **Soda** | | | | | |
| **1g WFR** | | | | | |
| <1g | 108 (93.1) | 7 (6.0)† | 94.0 | - | - |
| ≥1g | 0 (0.0)‡ | 1 (0.9) | | | |
| **15g WFR** | | | | | |
| <15g | 108 (93.1) | 7 (6.0)† | 94.0 | - | - |
| ≥15g | 0 (0.0)‡ | 1 (0.9) | | | |
| **Sugar with tea** | | | | | |
| **1g WFR** | | | | | |
| <1g | 70 (60.3) | 10 (8.6)† | 77.6 | 0.45 | 0.27, 0.63 |
| ≥1g | 16 (13.8)‡ | 20 (17.2) | | | |
| **15g WFR** | | | | | |
| <15g | 75 (64.7) | 13 (11.2)† | 79.3 | 0.45 | 0.26, 0.64 |
| ≥15g | 11 (9.5)‡ | 17 (14.7) | | | |
| **Juice** | | | | | |
| **1g WFR** | | | | | |
| <1g | 112 (96.5) | 4 (3.4)† | 96.5 | - | - |
| ≥1g | 0 (0)‡ | 0 (0) | | | |
| **15g WFR** | | | | | |
| <15g | 112 (96.5) | 4 (3.4)† | 96.5 | - | - |
| ≥15g | 0 (0)‡ | 0 (0) | | | |

*WFR=weighed food record; IVR=Interactive Voice Response; †False positive finding (type I error); ‡False negative finding (type II error)*

**SI Table 3**: IVR questionnaire, including skip logic and translation.

| Question number | Question | Recording label | Skip logic |
|---|---|---|---|
| | **Response options:**<br>If you ate, press 1<br>If you did not eat, press 3<br>If you do not know, press 8 | | |
| 1-40 | Questions for child dietary diversity | | |
| 41 | Now, I would like to ask you (mother), what you ate yesterday during day and night. Tell me everything that you ate at home or any other place like the neighbour's place even if they were small foods you ate. | Introduction message | |
| 42 | I am going to ask you the different types of food you ate, even those that were mixed with other foods. | Explanation message | |
| 43 | Yesterday, during day and night, did you eat or drink porridge, bread, rice, macaroni, millet, sorghum? | Grains | |
| 44 | Yesterday during day and night, did you eat pumpkin, carrots, orange fleshed sweet potatoes) | Vitamin A rich vegetables | |
| 45 | Yesterday during day and night, did you eat plantain, white sweet potatoes, yams, cassava, Irish potatoes | White roots / tubers | |
| 46 | Yesterday during day and night, did you eat dark green leafy vegetables like sukuma wiki, cassava leaves, pumpkin leaves, cowpea leaves, malakwang, spider leaf, okra and alayu | Dark green leafy vegetables | |
| 47 | Yesterday during day and night, did you eat any other vegetables like cabbage, tomatoes, and eggplants? | Other vegetables | |
| 48 | Yesterday, during day and night, did you eat ripe mangoes or ripe papaya | Vitamin A rich fruits | |
| 49 | Yesterday during day and night, did you eat, any other fruits like oranges, passion fruits, pineapple, soursop fruit, and watermelon? | Other fruits | |
| 50 | Yesterday during day or night, did you eat liver, kidney, or heart? | Organs | |
| 51 | Yesterday, during day or night, did you eat sausage, canned meat? | Sausage | |
| 52 | Yesterday during day and night, did you eat any other meat like goat meat, beef, pork, chicken, duck meat, mutton, big rat meat, squirrel, or any other wild meat? | Meat/poultry | |
| 53 | Yesterday during day and night, did you eat eggs? | Eggs | |
| 54 | Yesterday during day and night, did you eat fresh fish or dry fish? | Fish | |

| | | | |
|---|---|---|---|
| 55 | Yesterday, during day and night, did you eat beans, pigeon peas, peas, cow peas, green gram? | Legumes/ pulses | |
| 56 | Yesterday during day and night, did you eat or drink ground nut, oyster nuts, cashew, simsim? | Nuts/seeds | |
| 57 | Yesterday, during day and night, did you drink milk, cheese, sweetened milk or sour milk? | Milk, cheese | |
| 58 | Yesterday, during day and night, did you eat white ants or grasshopper? | White ants grasshoppers | |
| 59 | Yesterday during day and night, did you eat fish eggs? | Fish eggs | |
| 60 | Yesterday during day or night, did you eat sheabutter, sunflower oil, lard, groundnut oil | Oil, Fat | |
| 61 | Yesterday, during day or night, did you anything sweet like chocolate, sweet, ice cream, cake, biscuit? | Chocolate, sweets | |
| 62 | Yesterday, during day and night, did you eat fried cassava chips, fried cassava, irish chips, chapatti and mandazi? | Fried foods | |
| 63 | Yesterday during day and night did you drink juice, juice made from home or bought from the shop? | Juice | |
| 64 | Yesterday, during day and night, did you drinks like soda, rockboom or sting? | Soda | |
| 65 | Yesterday during day or night, did you take sweetened tea, sweetened coffee or any other sweetened or sweetened herbal drink? | Sweetened tea/coffee | |
| 66 | Yesterday, during day or night, did you drink other drink you like, like alcohol? | Other drinks | If NO(3)/DK(8), skip to 68 |
| 67 | If drunk, was it sweetened or not? | Other drinks sweetened | |
| 68 | Yesterday, during day or night, did you eat any other foods? | Other foods | |
| 69-88 | Questions on women's handwashing practices | | |
| 89 | This is the end. Thank you very much | End remarks | |

*IVR=Interactive Voice Response*
